# Exploring mental health staff’s views and experiences on supporting service users’ needs for romantic/intimate relationships: a qualitative systematic review

**DOI:** 10.1101/2025.04.01.25325034

**Authors:** Eunice Wu, Helen Killaspy, Sharon Eager, Aisling Smith O’Connor, Brynmor Lloyd-Evans

## Abstract

Mental health service users often voice a need for support regarding romantic/intimate relationships, yet staff face barriers in delivering that support. Given the importance of intimacy to well-being and recovery, this systematic review aimed to synthesise the available qualitative literature on mental health practitioners’ views and experiences of supporting people’s needs for romantic/intimate relationships. We conducted searches on four research publication databases. Quality of studies was assessed using the Joanna Briggs Institute Checklist for Qualitative Research, and results were summarised using meta-aggregation. Confidence in the findings was measured using the ConQual assessment tool. We identified 24 papers which met our inclusion criteria. Four synthesised findings were developed, namely 1) ideas and perceptions surrounding the intimacy needs of service users, 2) service provision at a personal level, 3) fitting intimacy needs into the therapeutic context and 4) service provision at an organisational level. Staff understood the importance of addressing intimacy needs, but voiced a need for improved knowledge, skills and support on how to have such conversations. Our findings can inform mental health policy change and support the development of interventions and guidelines that will enable staff to discuss with service users their needs regarding romantic/intimate relationships.

## Introduction

### Intimate relationships: definitions and importance

Love is a construct that has universal and enduring appeal. There have been many attempts to operationalise love, with one of the most prominent being Sternberg’s (1986) theory that love comprises passion, intimacy and commitment. While love may come in many forms, intimate relationships have been the subject of most scrutiny under a scientific lens. In this review, romantic/intimate relationships will be defined as relationships which are characterised by high physical, cognitive and emotional closeness (Schoebi & Randall, 2015), such as through the presence of romantic love, sexual relations or increased physical intimacy. Importantly, intimate relationships are only one part of the broader concept of sexuality, which is a “central aspect of being human throughout life” that encompasses all dimensions of sexual expression (World Health Organisation, 2016).

Being in a committed relationship has been linked with greater subjective well-being compared to singlehood (Soons & Liefbroer, 2008), although researchers suggest that relationship status does not solely explain this link (Kansky, 2018). Loneliness, which is increasingly recognised as a global public health problem (Department for Culture, Media & Sport, 2023; Surkalim et al., 2022) contributing to poor physical and mental health (Holt-Lunstad et al., 2015; Valtorta et al., 2016; White et al., 2021a), may also be mitigated by romantic/intimate relationships. Positive romantic/intimate relationships can help to address “intimate loneliness” – the absence of love, close companionship and confiding relationships – one of three proposed domains of loneliness (Cacioppo et al., 2015).

### Service users’ needs and barriers regarding intimate relationships

Among people with mental health problems, engaging in a committed romantic/intimate relationship has also been suggested to facilitate recovery (Boucher, Groleau & Whitley, 2016), as it provides a sense of normality and positive identity (Redmond, Harkin & Harrop, 2010). However, people with mental health conditions are less likely to enter a romantic/intimate relationship than those without mental illness for various reasons (Badcock et al., 2015; Braithwaite & Holt-Lunstad, 2017). These include the impact of the mental illness itself, for example, through the impairment of social and sexual functioning (de Jager et al., 2017), as well as decreased libido associated with some psychotropic medications (Montejo, Montejo & Baldwin, 2018), that may lead to an avoidance of intimacy (Mizock et al., 2019). In addition, mental illness is often associated with stigma (McCann et al., 2019); people with mental illness are rated significantly lower than those without mental illness on factors relating to “mate potential”, such as social status, sexual desirability and personality (Boysen, 2017). Experiencing such prejudices can negatively impact one’s self-esteem; one study reported that 41-81% of people with mental illness considered themselves to be “devalued” romantic partners (Wainberg et al., 2016). Additionally, people with severe mental illness are highly vulnerable to sexual and/or financial exploitation (Boysen & Isaacs, 2022; Kaul et al., 2024; Killaspy et al., 2017; Killaspy et al., 2019). For some service users, being in an institutional setting such as hospital or supported accommodation also reduces opportunities to develop romantic/intimate relationships (McCann, 2010). This may be due to program-level factors such as the lack of privacy and tight restrictions on guests, which may be compounded by often long lengths of stays (Tomlin et al., 2018).

### What does support currently look like?

Despite growing evidence for the need to support people with mental illness in finding and maintaining romantic/intimate relationships, as well as service users expressing a desire for support (Östman & Björkman, 2013), mental health staff often felt unprepared to address intimacy needs in their work (Urry et al., 2019).

This appears to contrast with recent shifts in the culture of mental health services in offering a more collaborative, personalise and recovery-oriented approach, that enables service users to have greater autonomy in building dignified and meaningful lives (Davidson et al., 2009). Within this perspective, intimacy as a fundamental expression of humanity should be seen as integral to one’s recovery process (de Jager et al., 2017). This has been reflected in certain healthcare policies around the world. In the UK, for instance, the Care Act 2014 identified that individuals are entitled to have support with personal relationships as these are an important component of well-being. National guidance also recommends that mental health and social cares staff should facilitate conversations with service users to talk to them about sexuality and relationships, as an essential prerequisite to offering any support to people to promote their sexual safety (Care Quality Commission, 2020).

Staff’s reluctance to discuss intimacy despite the shift towards recovery-oriented care may reflect the complexity of translating policy to practice and the lack of guidance on how to deliver this kind of support. Interventions targeting the reduction of loneliness and social isolation in adults with mental disorders have been developed, but the evidence is not robust, and there is typically not a focus on romantic/intimate relationships (Ma et al., 2020). More recently, a systematic review reported a lack of studies of interventions for people who were not already in established relationships (Caiada et al., 2024).

As a result, clinicians may feel deskilled in raising the subject of intimacy with service users (Dyer & das Nair, 2013), and clinicians may choose to only broach the subject if issues surrounding sexual vulnerability or risk are identified (Forrester-Jones, Dixon & Jaynes, 2023). Often, mental health practitioners expect service users to bring up the topic of romantic/intimate relationships or sex first (Quinn, Happell & Browne, 2011). However, service users report feeling distrust in services and apprehension about initiating such conversations, even if they desire support (White et al., 2021b; Eager et al., 2023).

### Objectives of the current study

The most recent systematic review on service users’ views on receiving support regarding intimate relationships was published by McCann and colleagues (2019), but to the authors’ knowledge, there has been no review on staff’s perspectives to date. This study aimed to synthesise the available qualitative literature on mental health practitioners’ views and experiences of supporting service users’ needs for romantic/intimate relationships as part of their provision of holistic care.

## Methods

### Protocol

The study protocol was prospectively registered in the PROSPERO database on 10^th^ of April, 2024 (CRD42024534633). The protocol was drafted in compliance with the PRISMA 2020 Checklist (Page et al., 2021).

### Eligibility Criteria

#### Phenomena of interest

We conducted a systematic review of qualitative studies, focused on understanding mental health staff’s views and experiences of supporting the need for romantic love, sexual relations or increased physical intimacy in people receiving mental health care.

#### Population

We included studies with any staff who provide care and support to people using adult mental health services. Studies involving mental health staff who specialised in addressing romantic/intimate relationship needs, such as relationship counsellors, sex therapists and psychotherapists were excluded.

#### Context

Studies that were conducted in any inpatient or community based mental health service provided by public (e.g. NHS or Local Authority), independent/private or voluntary sectors were eligible for inclusion.

#### Types of studies

Eligible studies were peer-reviewed and non-peer reviewed studies which used qualitative designs. These included, but were not limited to, studies using semi-structured interviews, topic guide facilitated interviews, surveys with free text responses and/or focus groups. Studies using any approach to qualitative data analysis (such as thematic analysis) were eligible for inclusion. Qualitative findings from mixed methods studies were also included.

No geographical restrictions or limitations on the date of publication were imposed. In a deviation from our protocol, we did not include an English-language restriction; studies published in any language were eligible for inclusion.

### Search Strategy

The search strategy involved three stages: 1) searching four electronic bibliographic databases, 2) searching two sources of grey literature and 3) searching via forward and backward citation searching. Search terms for the academic databases were developed using the PICOS framework following consultation with a subject librarian. The search terms referenced four concepts: 1) romantic/intimate relationship needs, 2) mental health, 3) staff and 4) qualitative research. Terms that pertained to the same concept were joined by the Boolean operator “OR”. Subsequently, all concepts were linked by the Boolean operator “AND”. This search strategy was initially created within MEDLINE and adapted to PsycINFO, Web of Science and CINAHL using database-specific subject headings where applicable (Appendix A). A total of four databases were consulted. Searches for MEDLINE and PsycINFO were conducted via OVID on 21^st^ April 2024, while Web of Science (via Core Collection) and CINAHL (via EBSCOHost) were searched on 22^nd^ April 2024. We conducted a grey literature search on 10^th^ May 2024 through Google Scholar and ProQuest Dissertations and Theses Global. Only the first 10 pages of results were screened for each source of grey literature to ensure relevance. After screening, the reference lists for all included studies were manually reviewed for additional relevant studies. Studies which have cited the included studies were manually reviewed on 15^th^ January 2025.

Studies retrieved from the search were downloaded and exported into the Covidence systematic reviewing software package (www.covidence.org) and were deduplicated. One reviewer (EW) screened the titles and abstracts of all potential studies to see if they met the inclusion criteria outlined above. A second independent reviewer (ASOC) screened 10% of the studies to check for inter-rater reliability. Any discrepancies between the reviewers were resolved through discussion. Full texts of retained studies were then retrieved and double screened by the two independent reviewers (EW, ASOC). Studies excluded at this stage were coded with the reason for exclusion.

### Assessment of methodological quality

Following inclusion in the review, one reviewer (EW) assessed the methodological quality of eligible studies using The Joanna Briggs Institute (JBI) Checklist for Qualitative Research (Lockwood, Munn & Porritt, 2015), a critical appraisal instrument (Appendix B). Each study was checked against a set of standardised guidelines, in which “yes”, “no”, “unclear” or “not applicable” were selected based on 10 criteria. Studies scoring >7 were considered high quality, >5 were considered medium quality and <5 as low quality. Studies were not excluded on grounds of low quality.

### Data extraction and synthesis

Data from included studies were extracted and synthesised by one reviewer (EW), then checked by a second reviewer (ASOC) using a predetermined standardised data extraction table. Information on the authors, year of publication, country of origin, study design, phenomena of interest, setting, and participant characteristics were extracted. We also included a column detailing whether studies reported any patient and public involvement (PPI) input. This was added considering the increasing recognition of lived experience expertise (Ocloo & Matthews, 2016). Discrepancies between the two reviewers were examined by a third independent reviewer (BLE) and resolved through discussion.

Qualitative research findings were pooled using the meta-aggregative approach to narrative synthesis (Lockwood, Munn & Porritt, 2015). Findings from all included studies were extracted with an accompanying illustrative quote and allocated one of three levels of plausibility: unequivocal, equivocal or unsupported (Appendix C). Unsupported findings were not included in the synthesis. Findings were then combined inductively and iteratively to form categories for findings, with at least two findings included per category. Two or more like categories were then combined to form synthesised findings.

The ConQual approach was used to determine how much confidence to place in our synthesised findings (Munn et al., 2014). Synthesised findings were rated on a scale of “high”, “moderate”, “low” or “very low” based on the contributing studies’ dependability and credibility. The full protocol for ConQual can be found in Appendix D. In a deviation from our protocol, we decided to use the ConQual approach to assess confidence in our review findings instead of GRADE-CERQual, as this integrated better with the meta-aggregative approach to narrative synthesis.

### Reflexive Statement

Meta-aggregation, which involves extracting and combining findings from primary papers rather than reanalysing results to develop new, interpretative findings, was used to minimise any undue subjectivity in conducting the review. However, we recognise the analytic process of developing categories and synthesised findings requires decisions from the researchers which may be influenced by our positionality. Four of the authors are female and all are educated to at least post-graduate level. The team is comprised of researchers with a range of relevant academic and clinical experience in psychiatry and social work. All authors contributed to the analytic process and reviewed draft categories and synthesised findings.

## Results

### Study Inclusion

The search on bibliographic databases returned 8127 records. A total of 5844 records were screened following deduplication, from which 47 full-texts were retrieved for review and 17 papers were deemed eligible for inclusion. Exclusion reasons for studies excluded at full-text screening can be found in Appendix E. Our grey literature and backward and forward citation tracking searches yielded seven additional studies. A total of 24 papers reporting on 20 studies were included in the review. See figure 1 for the PRISMA flowchart of the search and study selection process.

**Figure 1.**
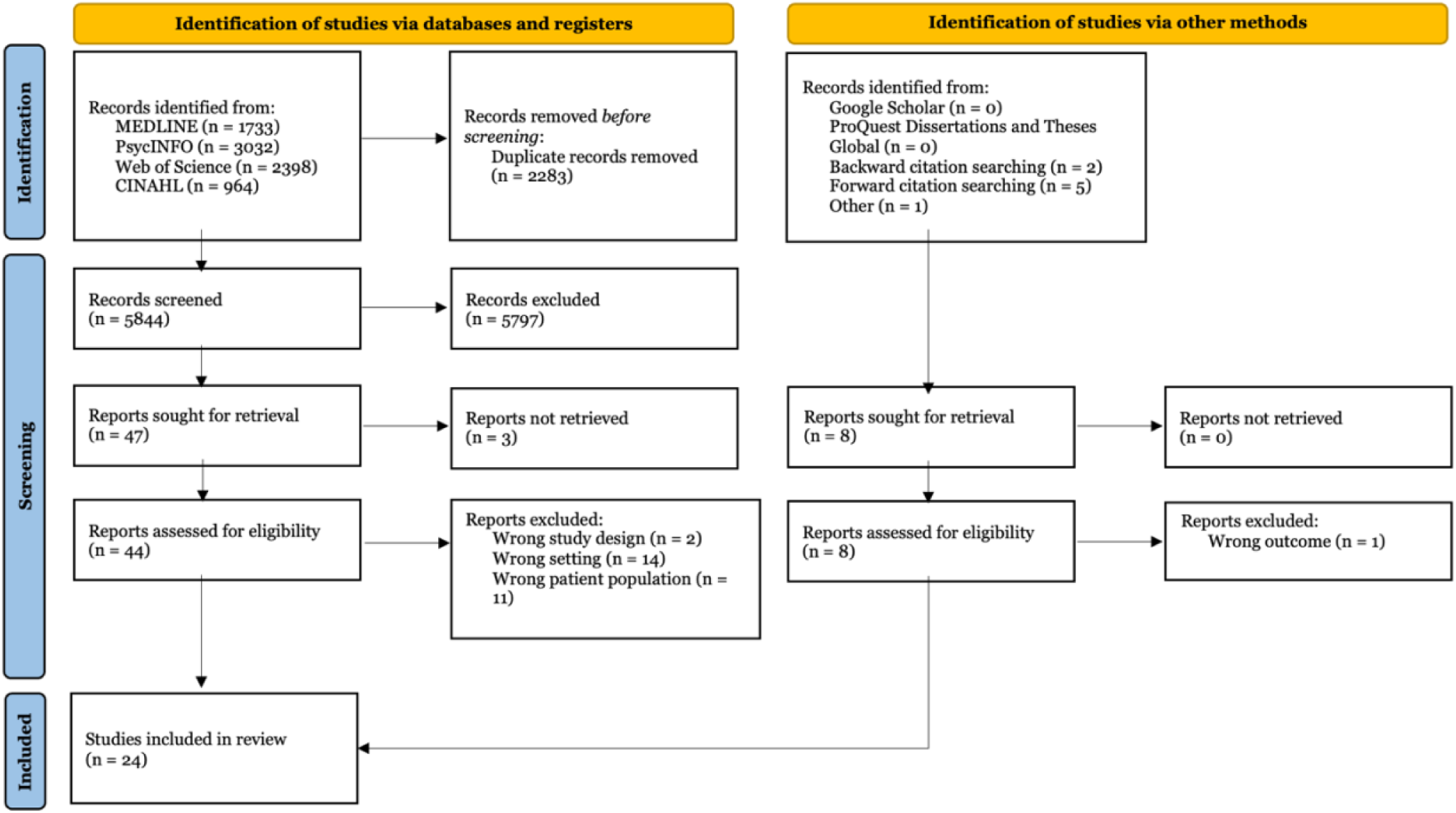
**PRISMA flowchart of the search and study selection process**

### Characteristics of included studies

Table 1 summarises the characteristics of included studies. Fourteen studies were published in the past five years. Studies were predominantly conducted in high-income countries, such as Australia (n=9) and United Kingdom (n=5). Sample sizes ranged from 5 to 219. Studies captured the perspectives from a range of mental health staff, including psychologists, psychiatrists, mental health nurses, social workers and other staff. Urry, Chur-Hansen & Khaw (2019) included three participants working specifically in sexuality or sexual health, however we elected to include the study as these participants represented a small minority of their whole participant sample (n=22). Most studies collected data through semi-structured interviews, and the remainder employed the use of focus groups, surveys with free text responses and collaborative reflection. Collaborative reflection was described as a process during which participants were asked to reflect on the results of a previous study. Analytic approaches used in the included studies were thematic analysis, case study analysis, content analysis and centroid factor analysis. Only three studies reported the involvement of people with lived experience in the research process.

**Table 1.**
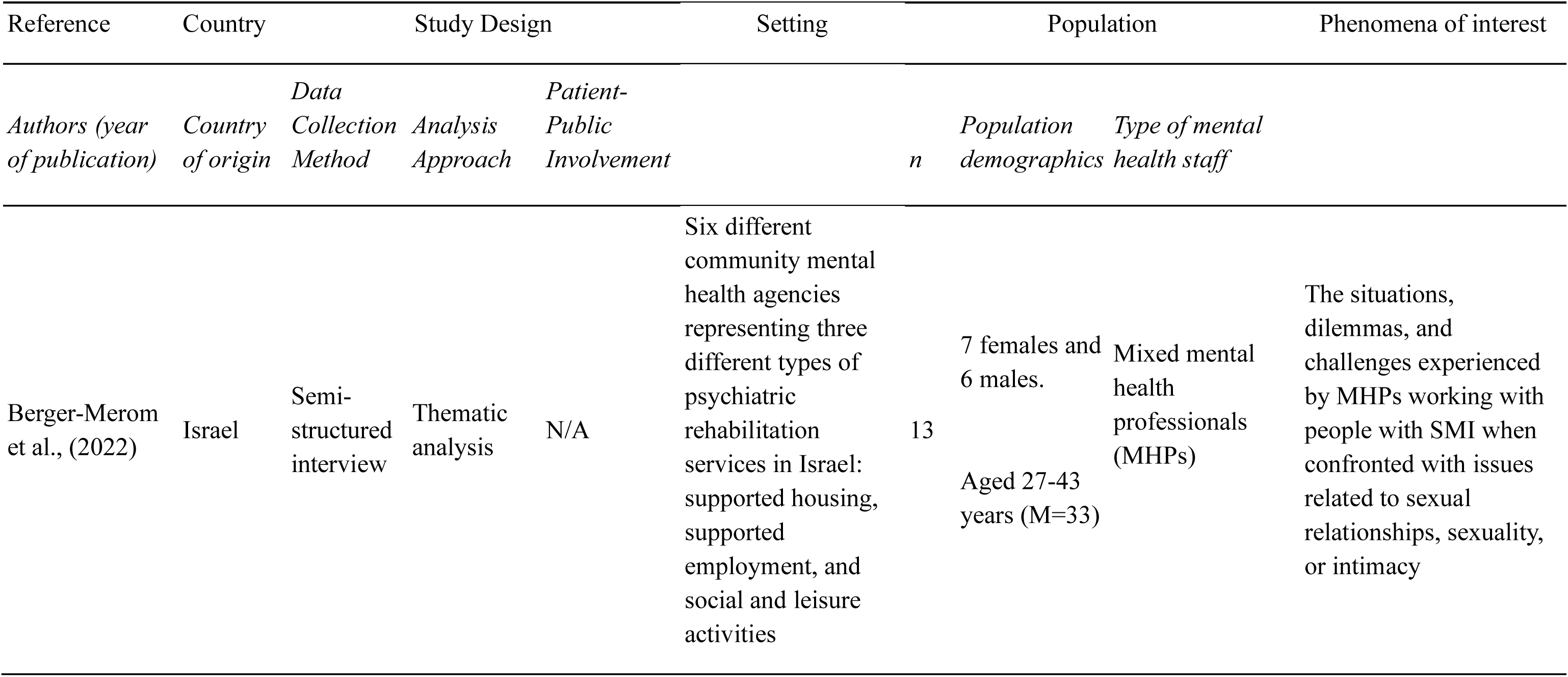
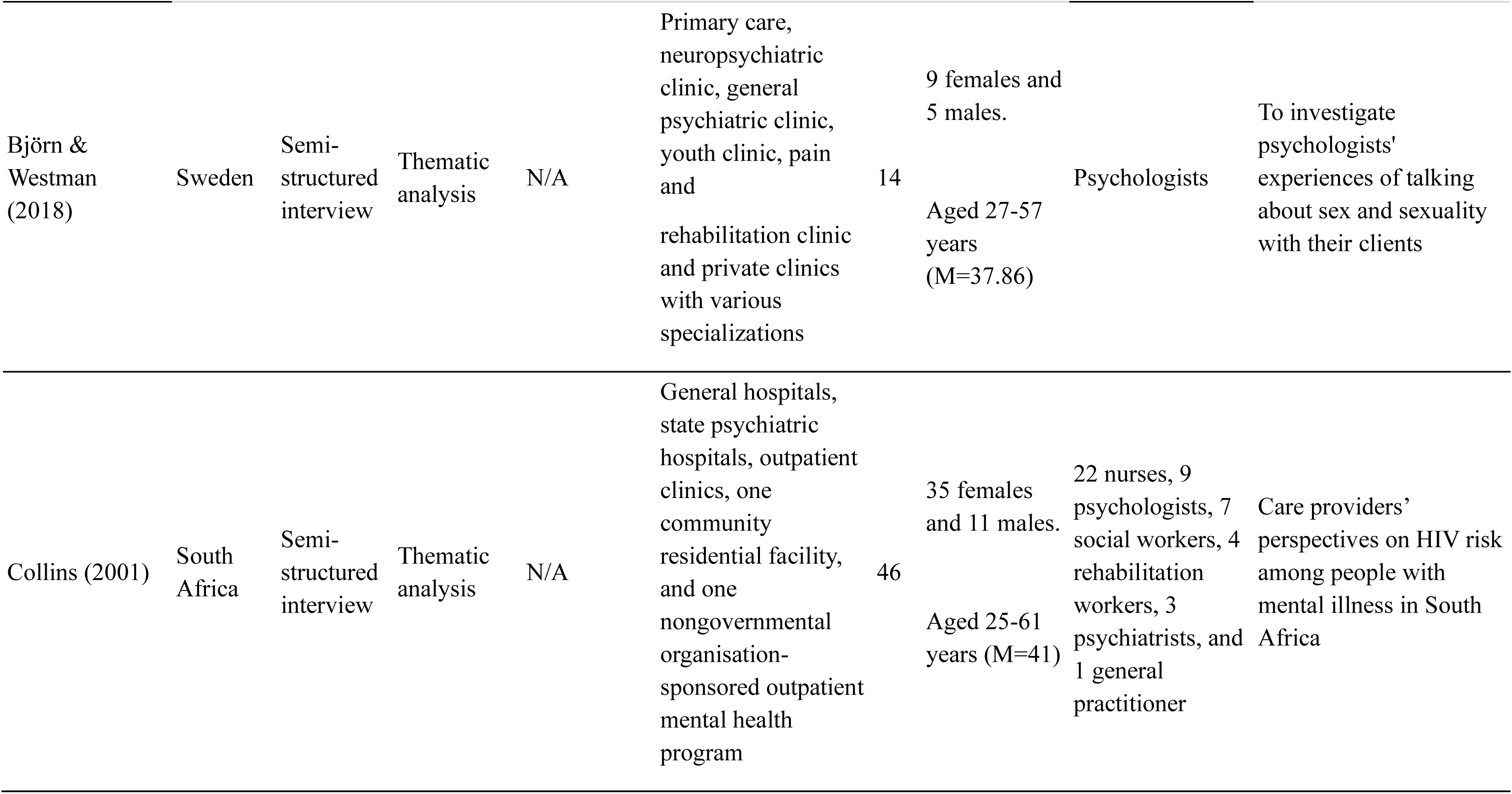
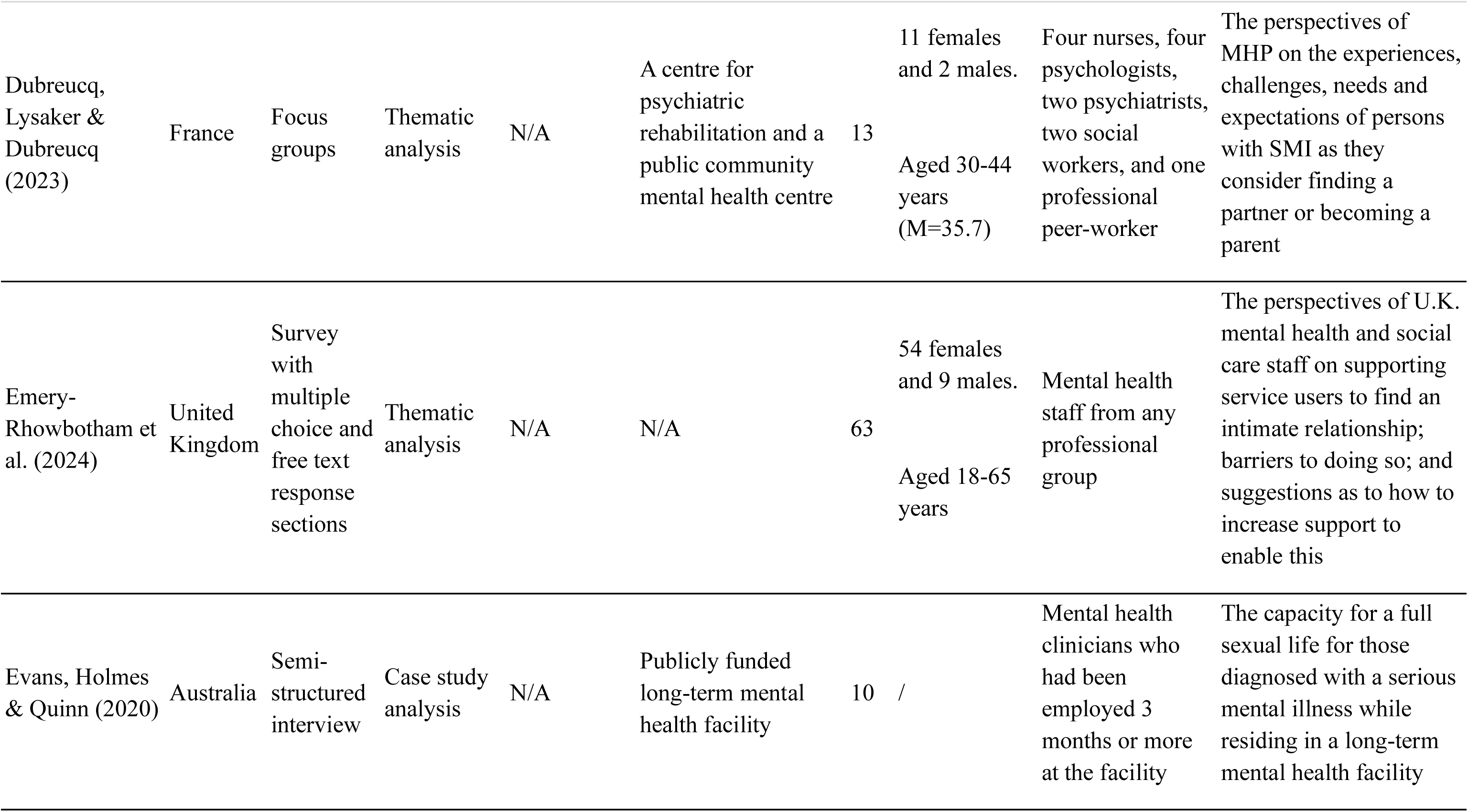
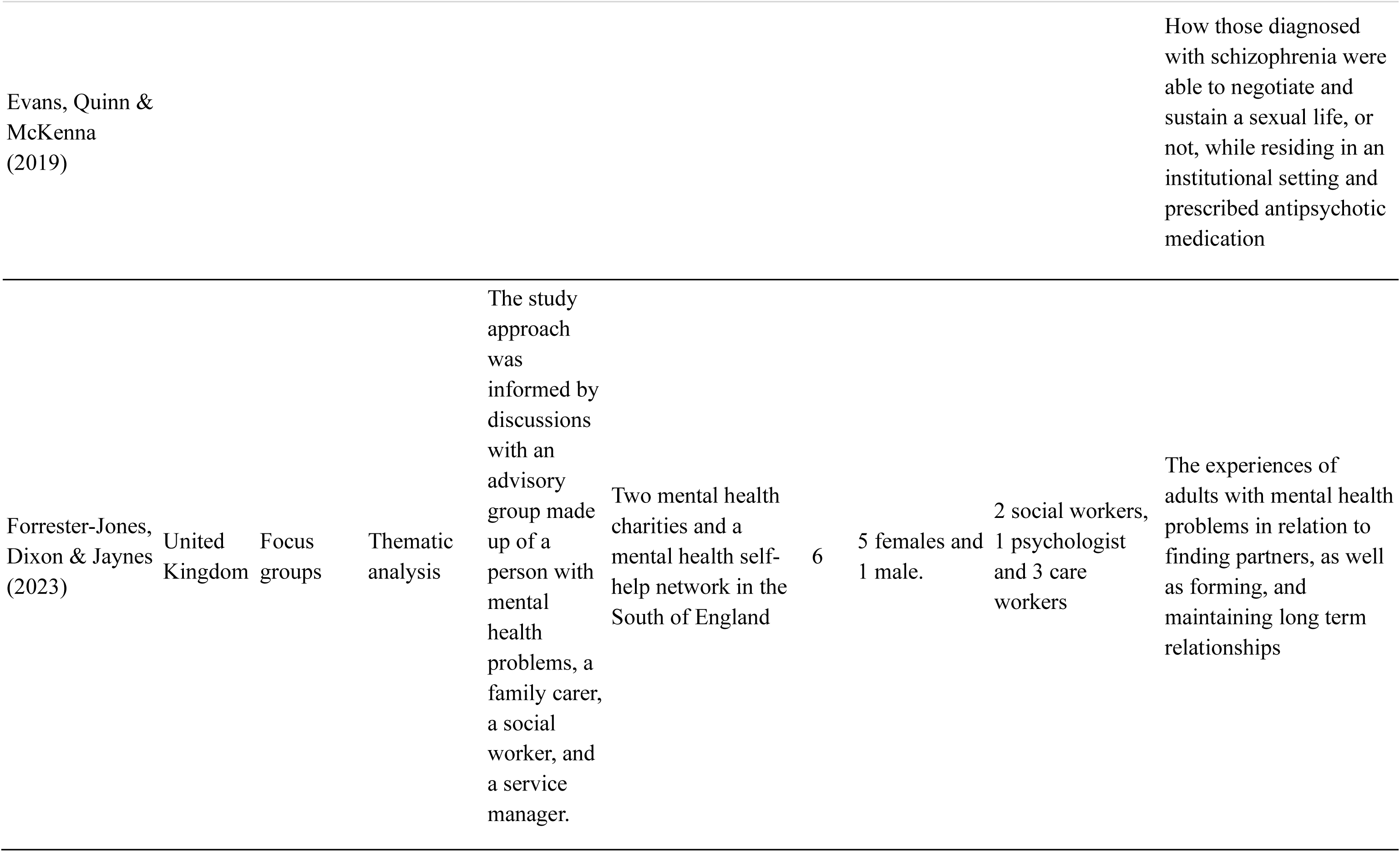
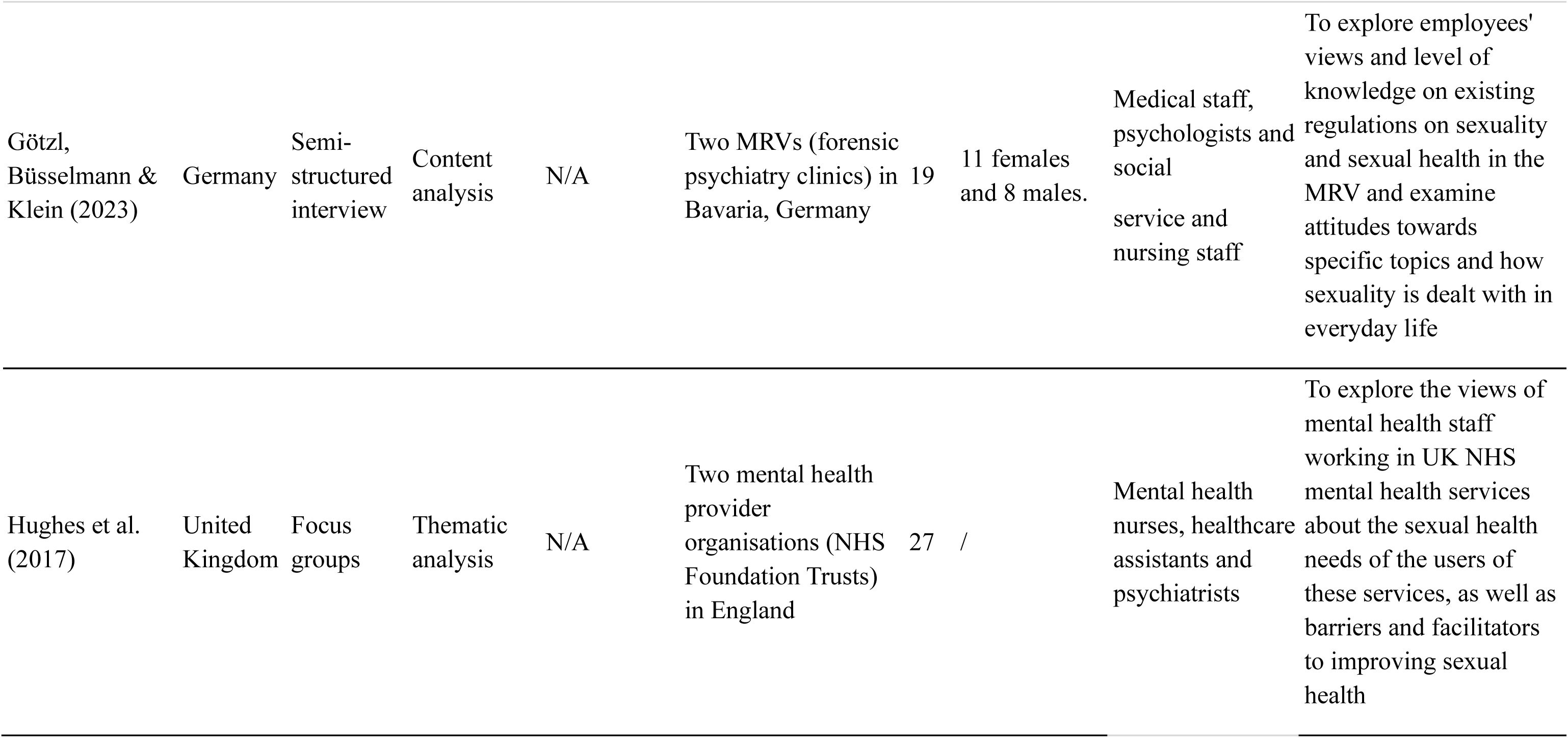
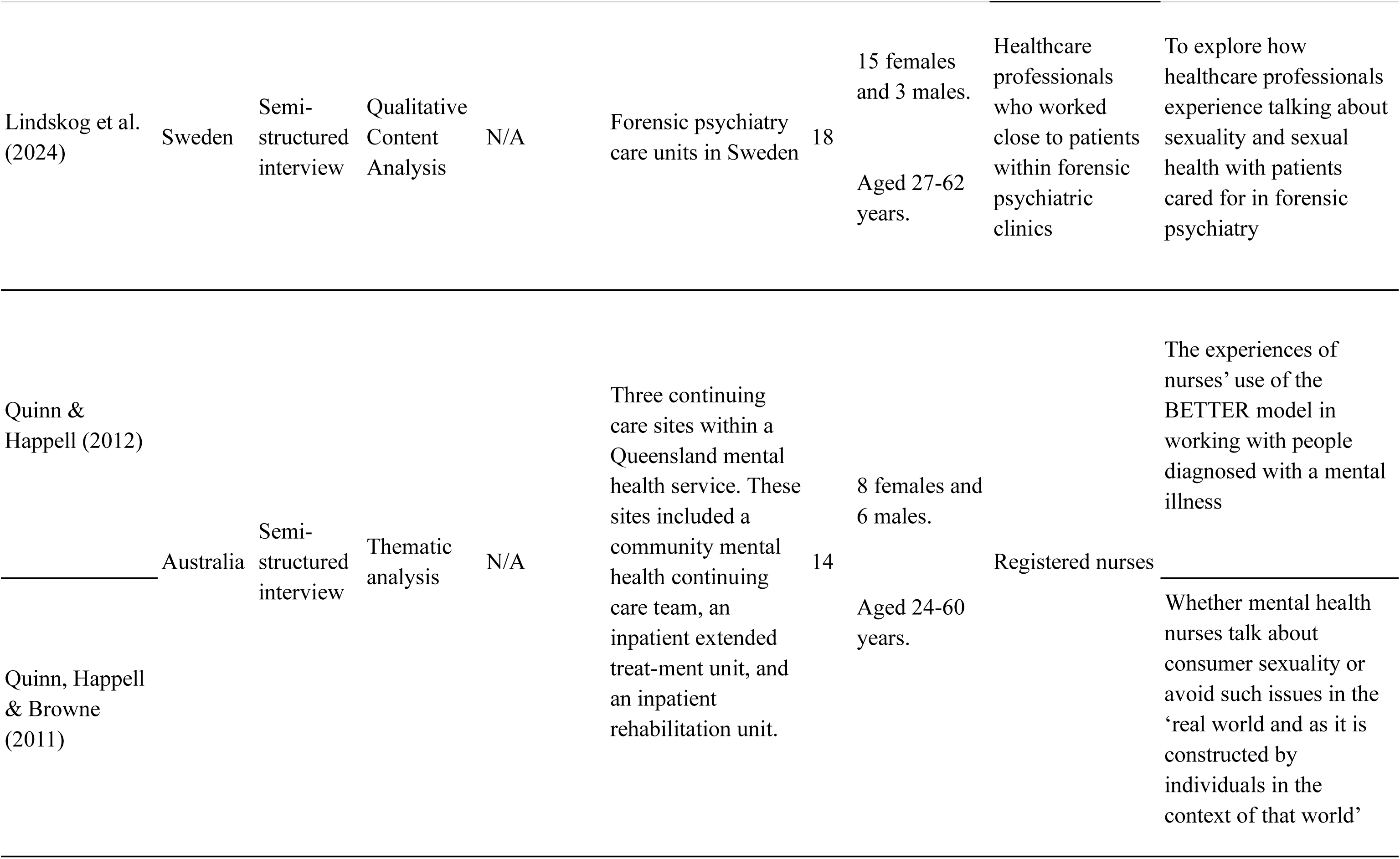
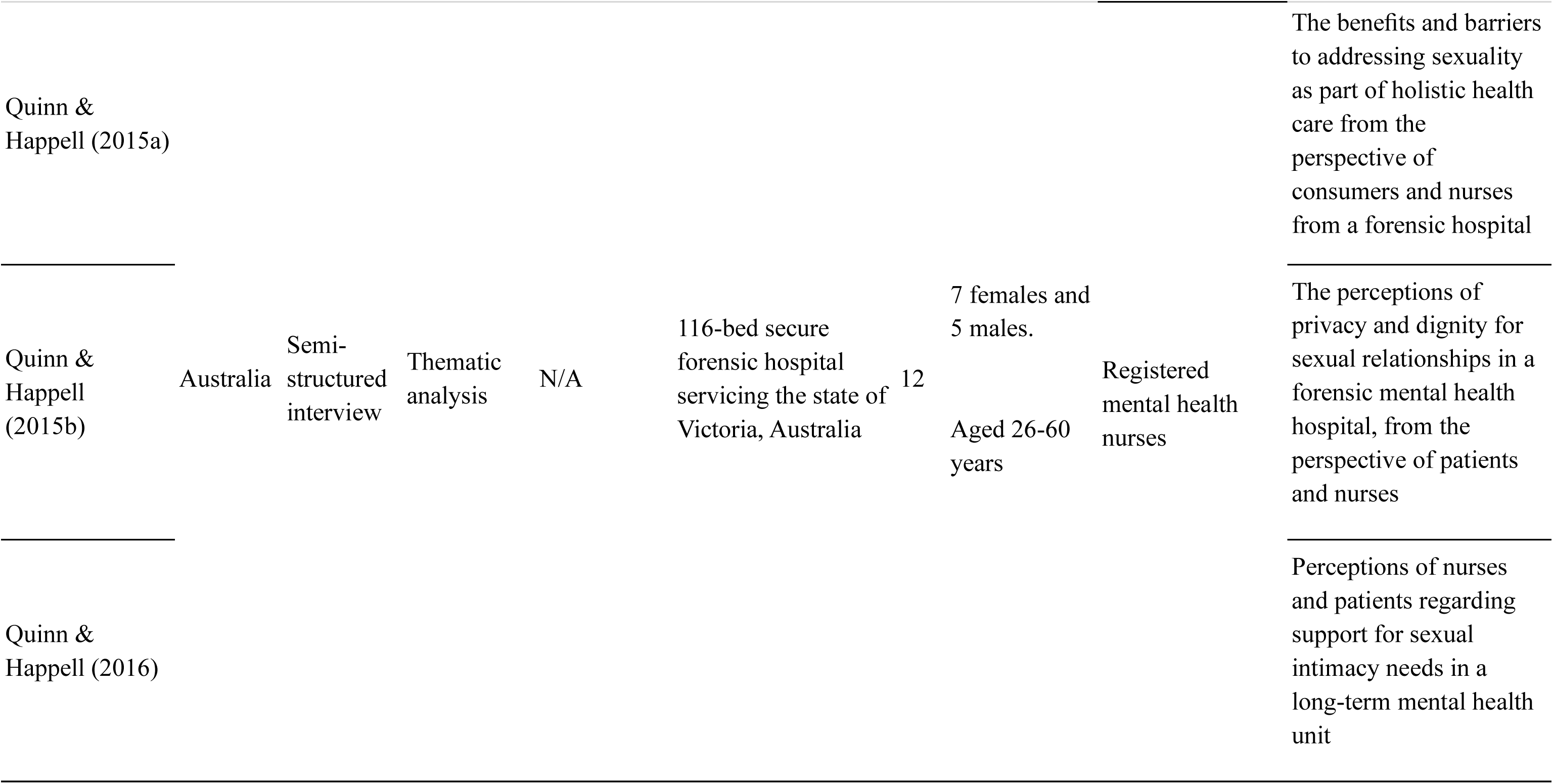
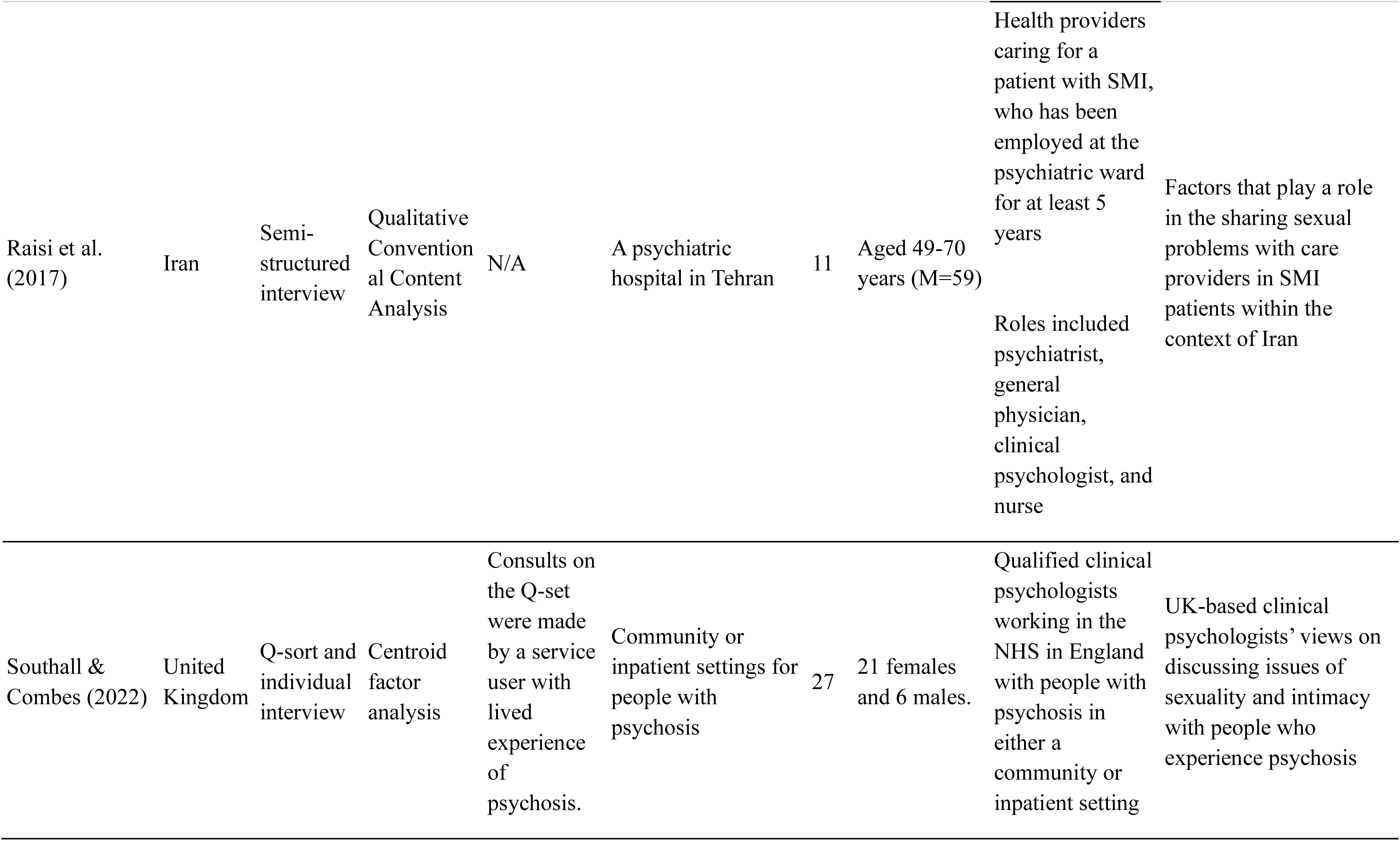
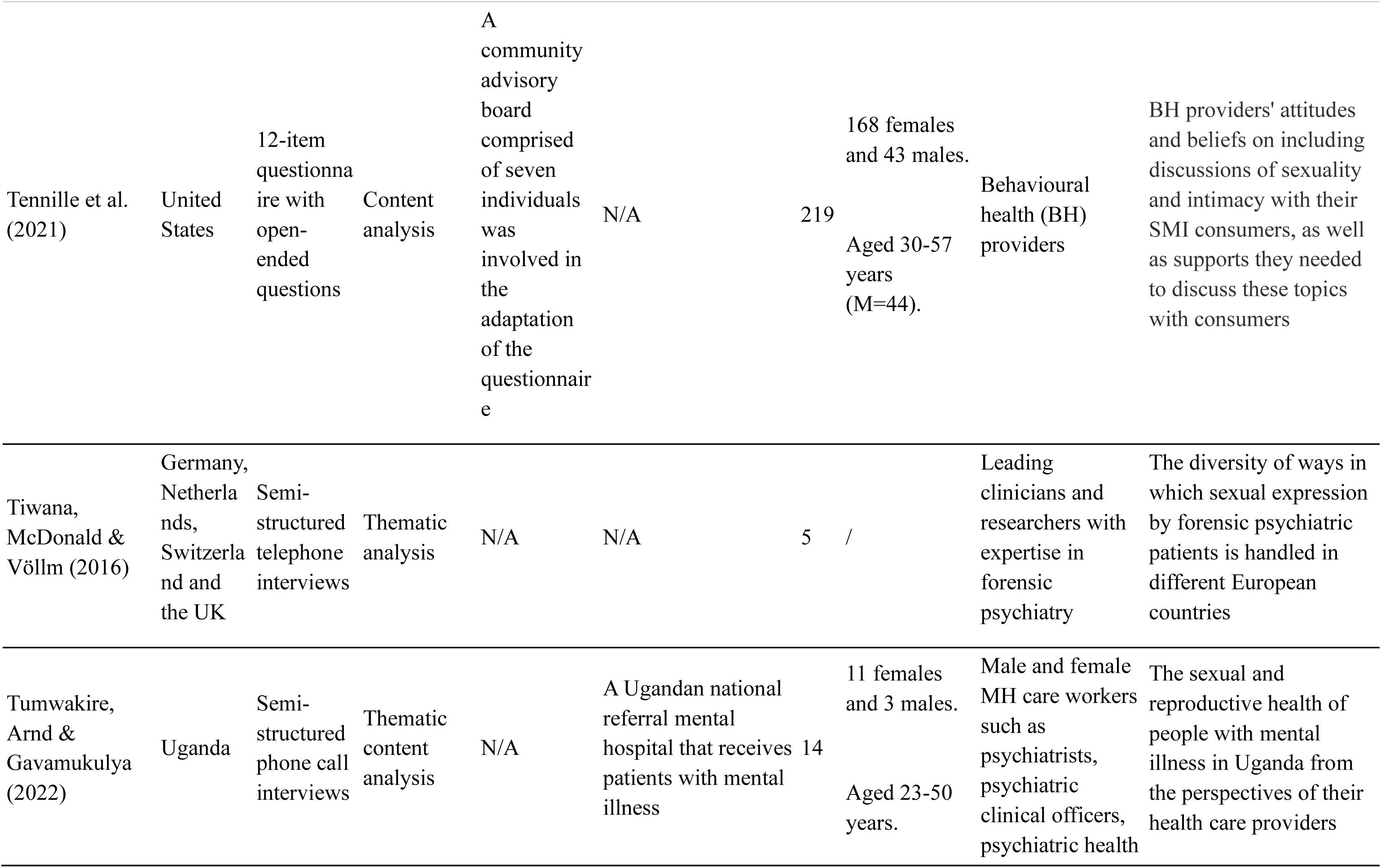
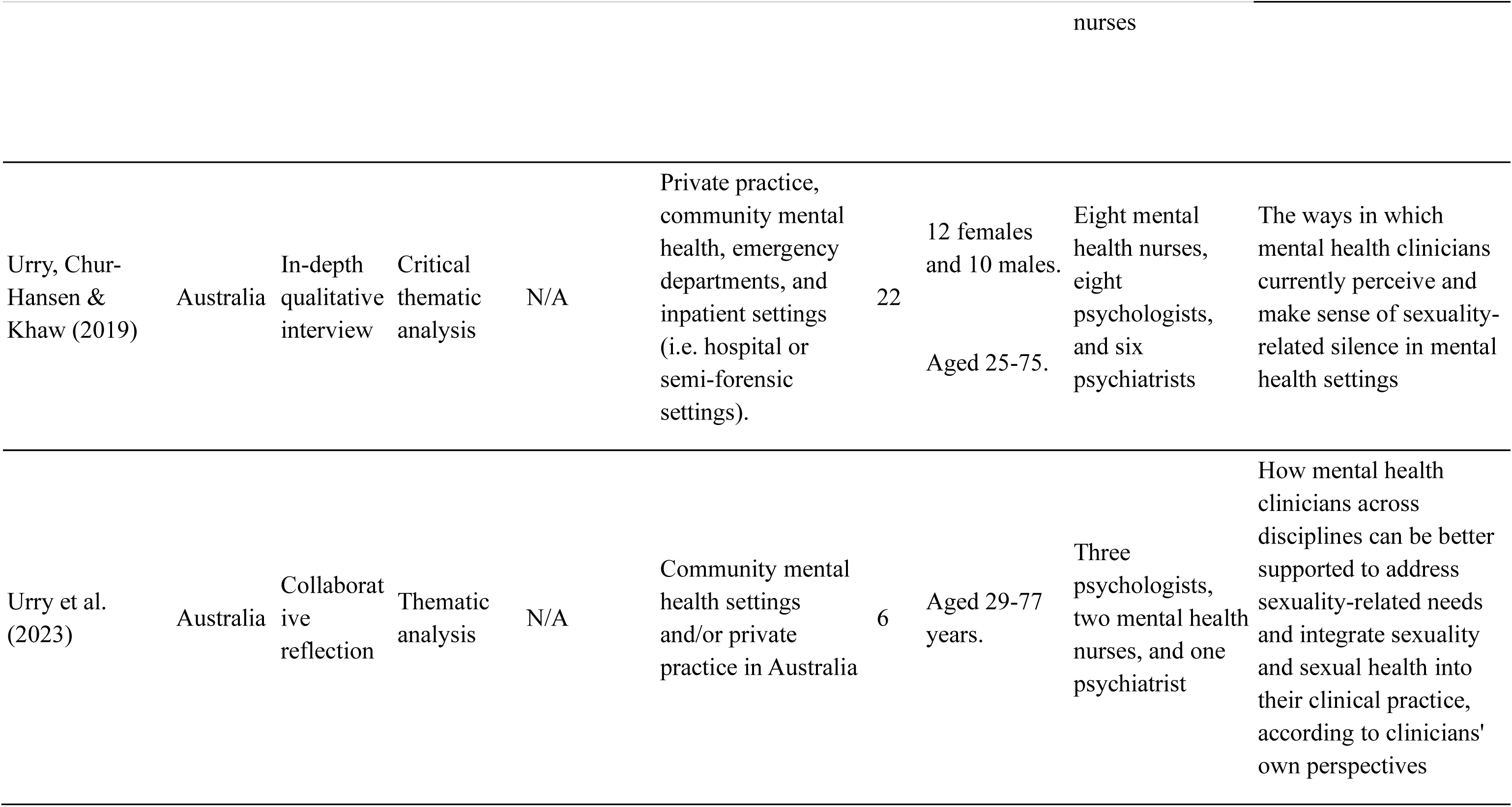
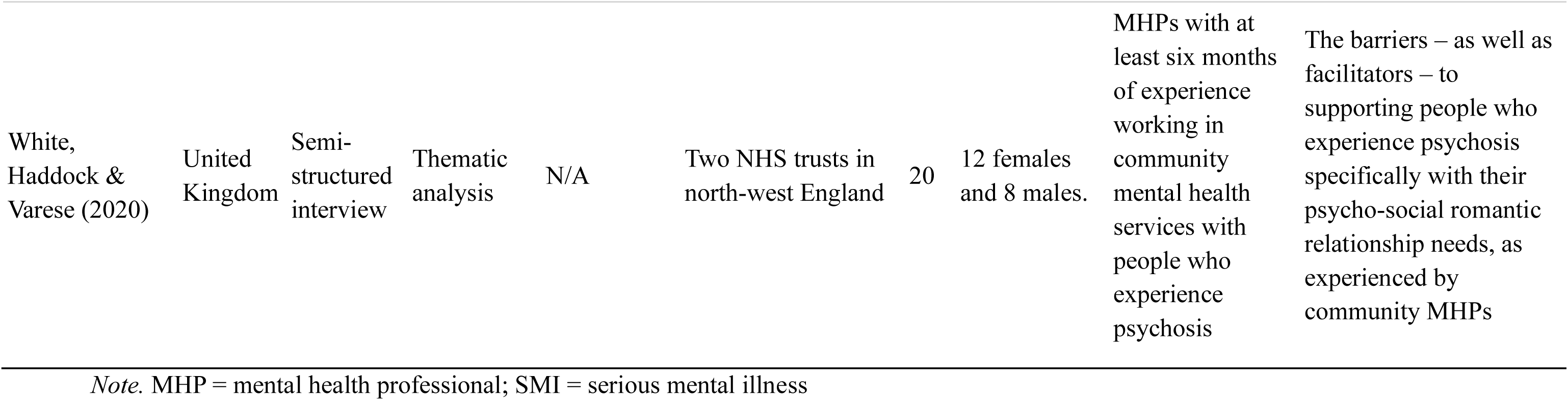
Characteristics of included studies.

### Methodological Quality

Table 2 contains the quality appraisal of all studies. All but one study scored at least 7, indicating generally high methodological quality. The remaining study achieved medium quality. Nineteen out of 24 papers failed to either provide a statement locating the researcher culturally or theoretically (Q6), or mention the influence of the researcher on the research and vice versa (Q7).

**Table 2.**
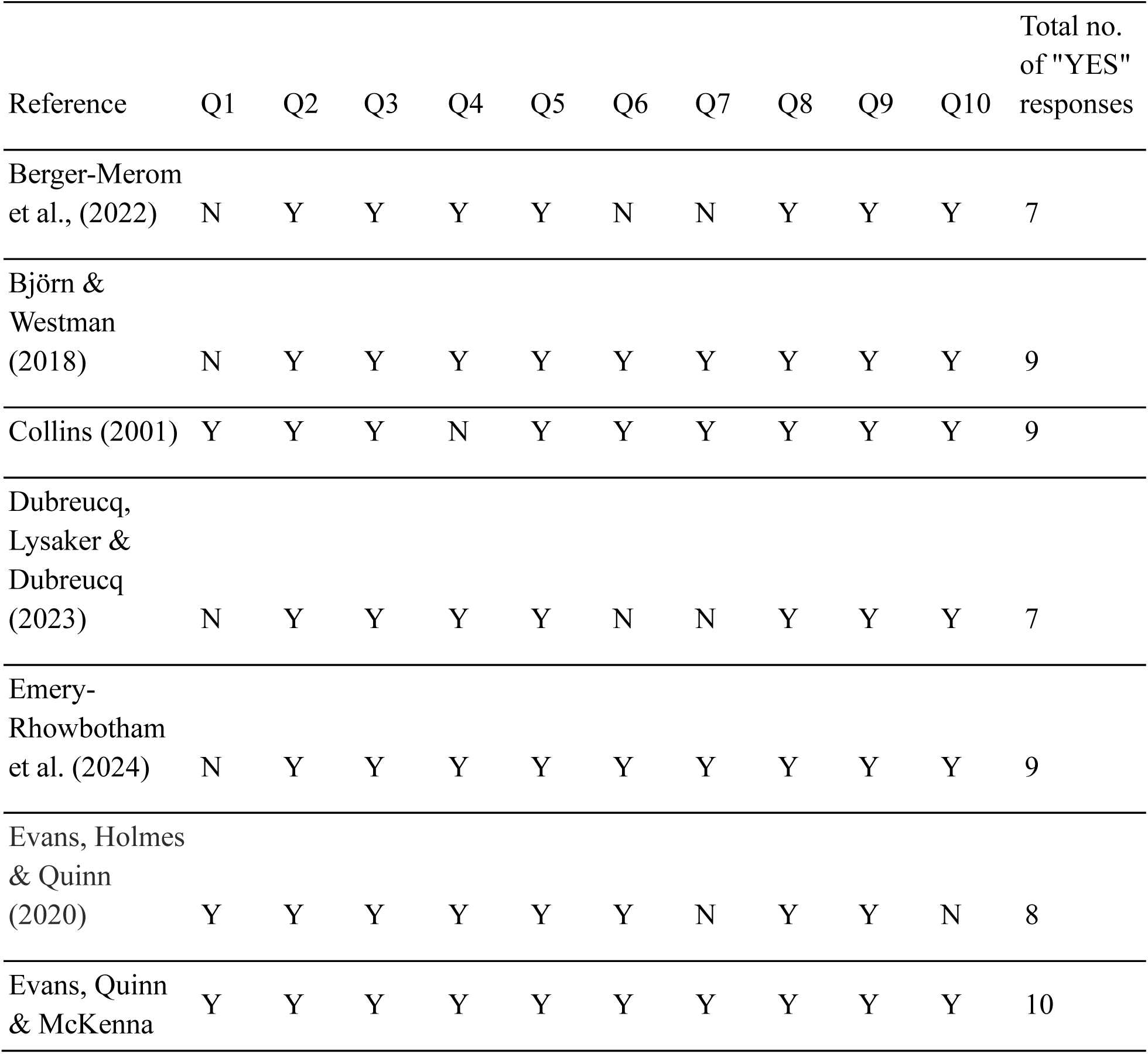
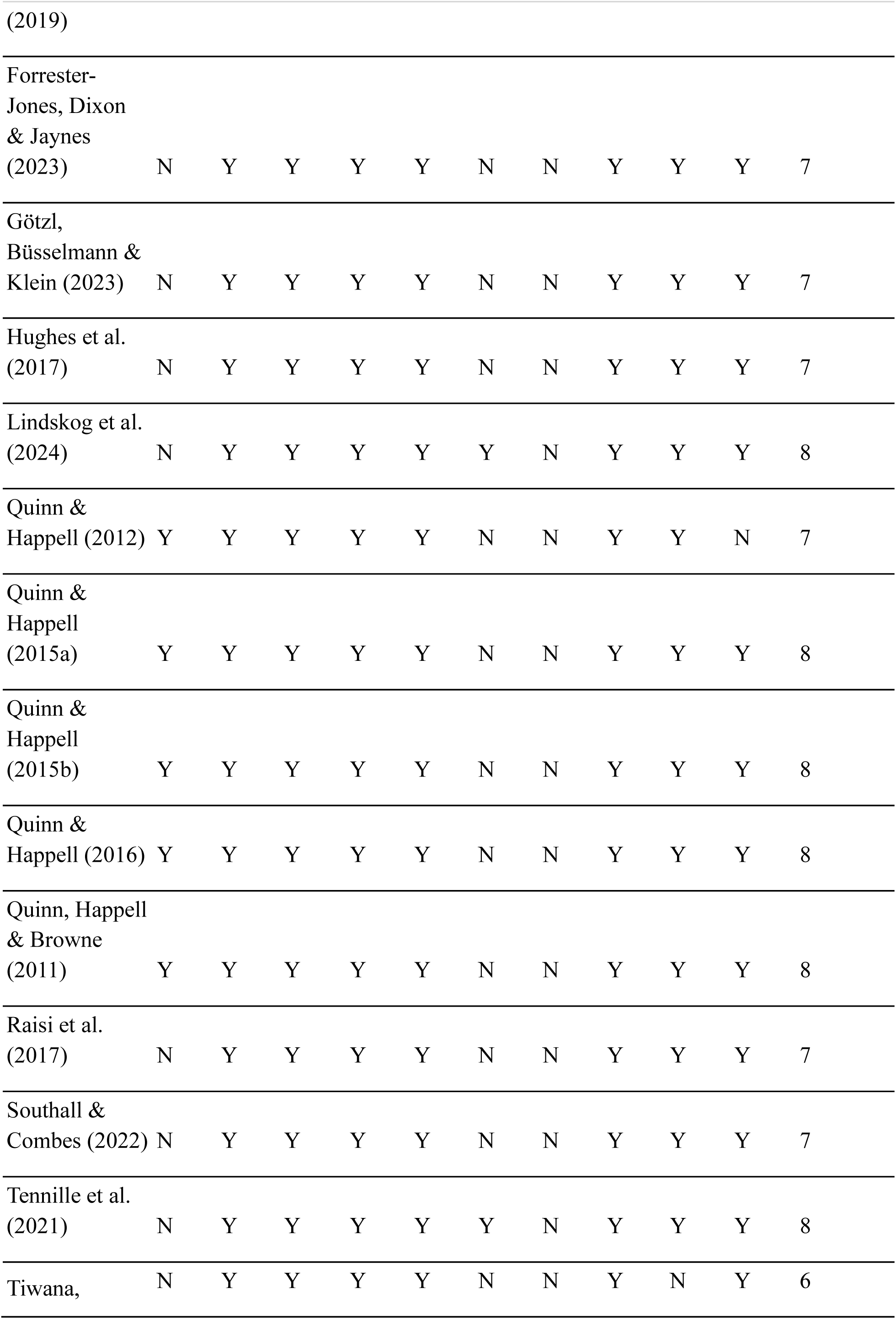
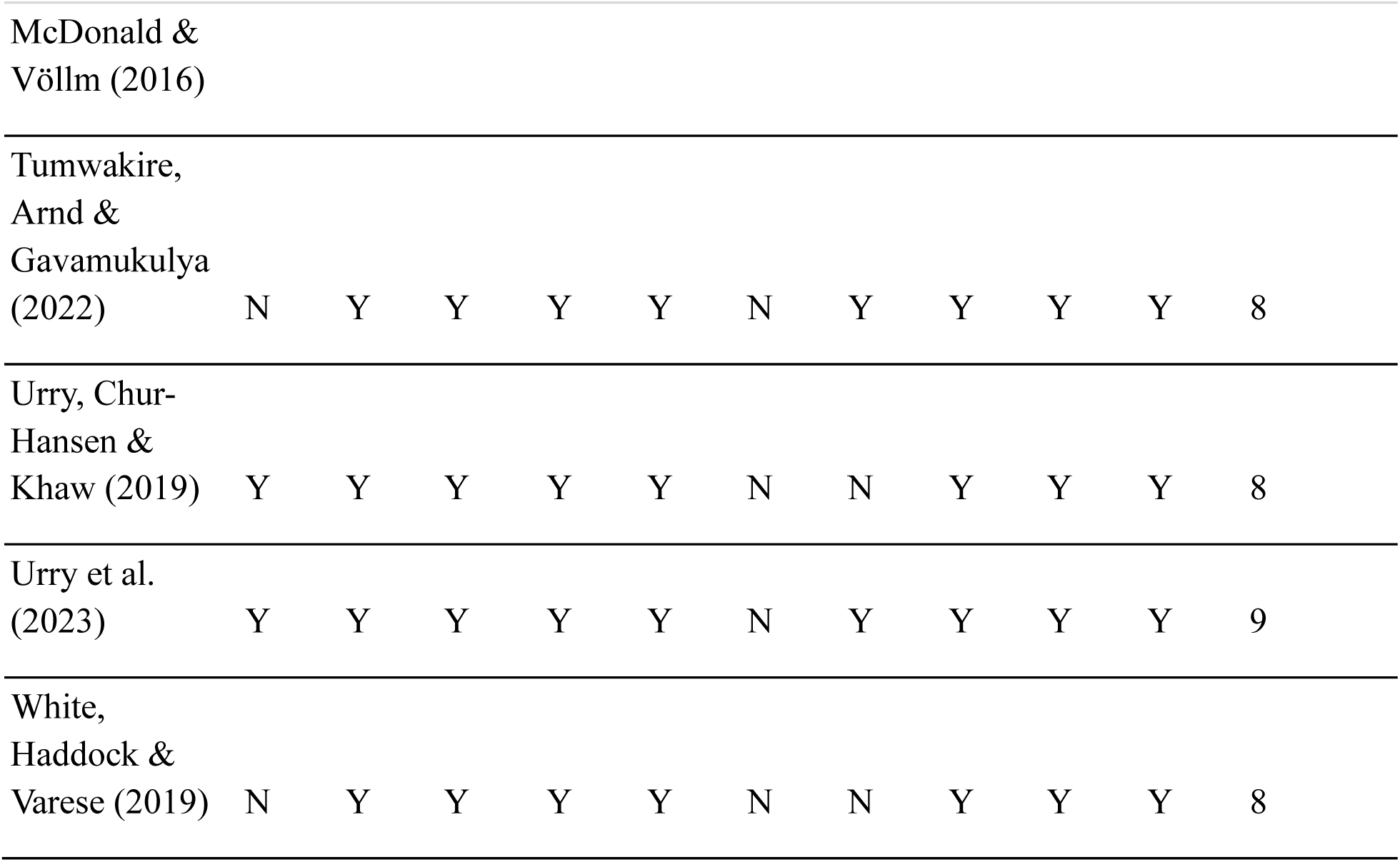
Methodological quality of included studies.

### Synthesised findings

We developed four synthesised findings: 1) ideas and perceptions surrounding the intimacy needs of service users; 2) service provision at a personal level; 3) fitting intimacy needs into the therapeutic context; and 4) service provision at an organisational level. The synthesised findings consisted of 163 findings grouped into 18 categories (Appendix F). A total of 147 findings were determined as “unequivocal”, 11 were “equivocal”. Six were “unsupported” which resulted in their exclusion from the analysis. Table 3 contains a summary of the synthesised findings, as well as the ConQual assessment of confidence in each finding. Confidence in one synthesised finding was rated as “moderate”, while three were rated as “low” due to the dependability and credibility of the studies contributing to the synthesised finding.

**Table 3.**
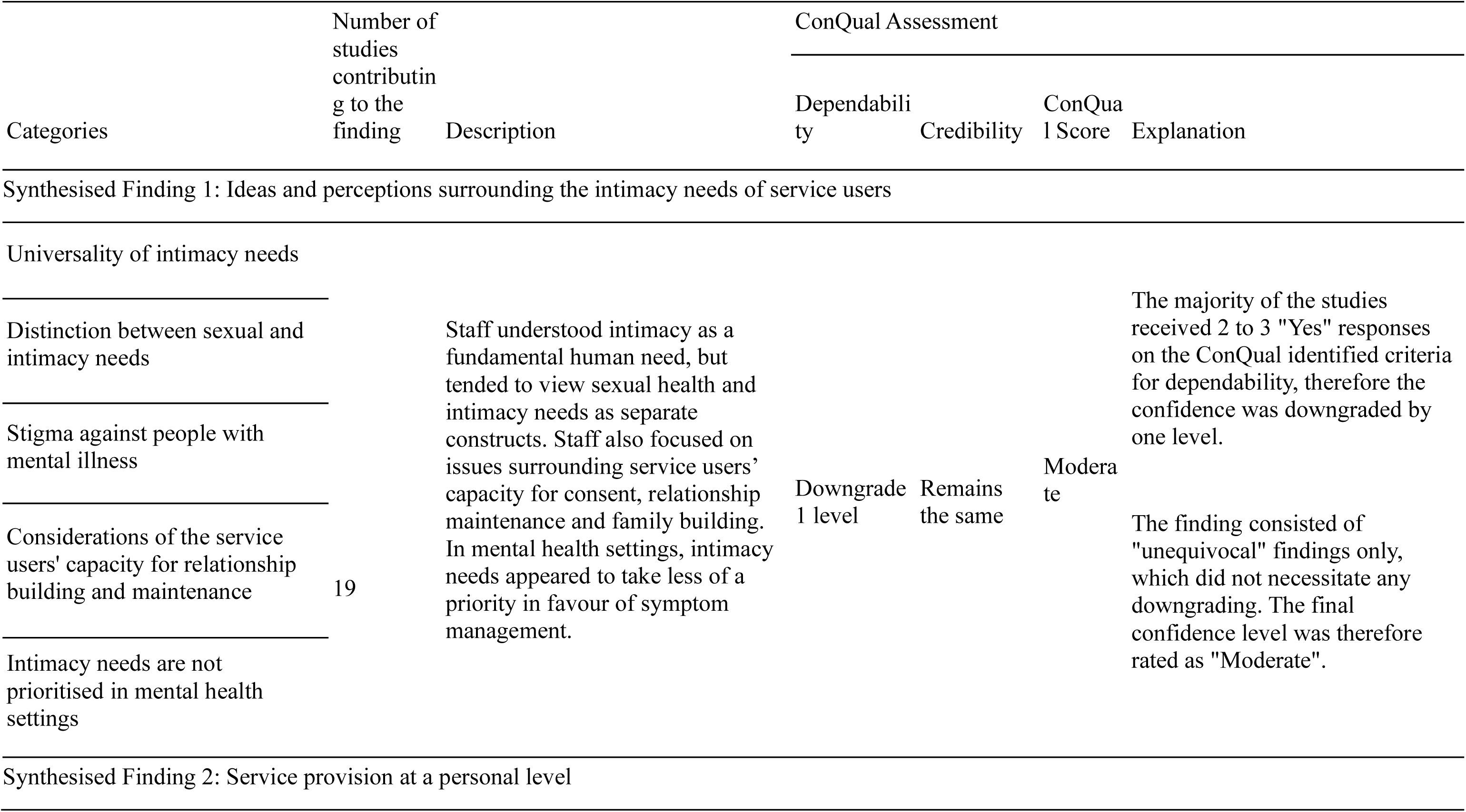
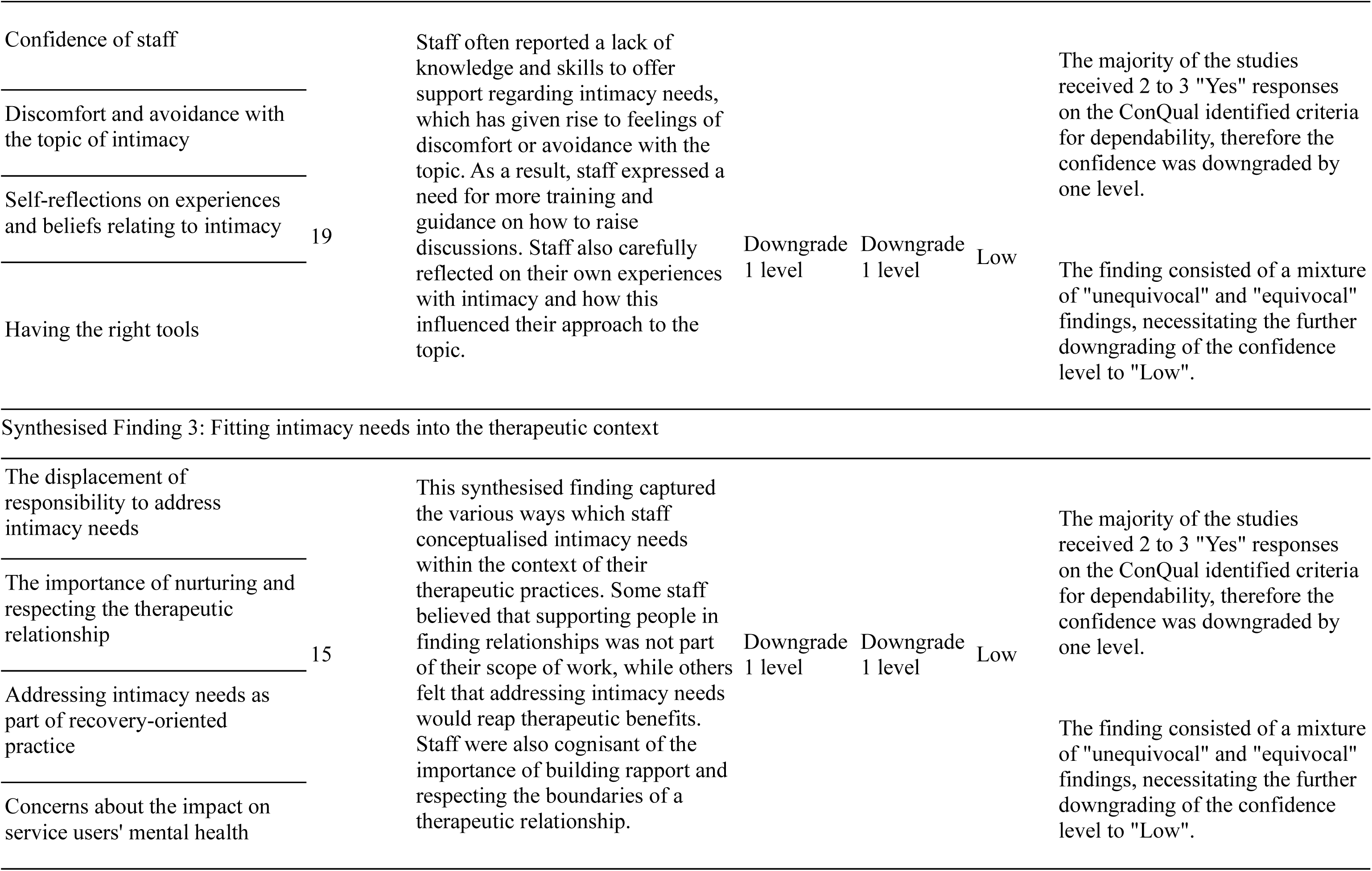
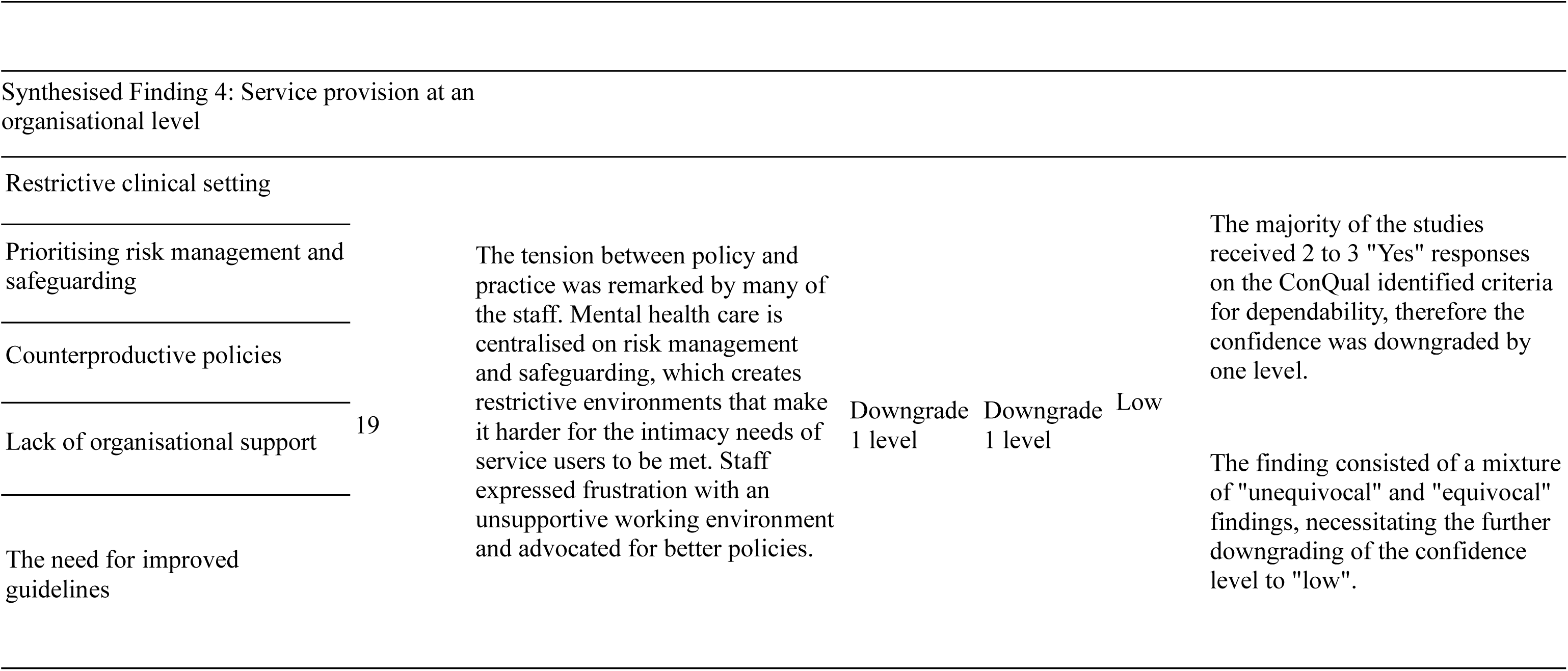
Summary of synthesised findings and accompanying ConQual assessment.

## 1. Ideas and perceptions surrounding the intimacy needs of service users

Staff understood intimacy as a fundamental human need, however they noted that stigma may impede service users’ success in finding and maintaining romantic/intimate relationships. When considering service users’ intimacy needs, staff tended to view sexual health and intimacy needs as separate constructs. Staff also focused on issues surrounding service users’ capacity for consent, relationship maintenance and family building. In mental health settings, intimacy needs appeared to take less of a priority in favour of symptom management. This synthesised finding received a ConQual score of “moderate” and was derived from 33 findings which were divided into five categories.

### 1.1 Universality of intimacy needs

Most staff felt that intimacy and sexual expression were a fundamental human right. They acknowledged that mental illness did not automatically equate to asexuality (Tumwakire, Arnd & Gavamukulya, 2022), and that the relevancy of intimacy in people’s lives warranted addressing in a clinical setting (Southall & Combes, 2022).

> *“We should know that being mentally sick doesn’t take away your sexual feelings, it doesn’t. These are normal women, these are normal men.” (Tumwakire, Arnd & Gavamukulya, 2022)*

### 1.2 Distinction between sexual and intimacy needs

Staff conceptualised sexual health, such as contraception and HIV prevention, differently from broader aspects of sexuality such as intimacy, pleasure and relationships. Education programs or discussions were sometimes limited to sexual health issues (Urry, Chur-Hansen & Khaw, 2019). One study revealed that goals tied to romantic/intimate relationships were seen by staff as legitimate recovery goals, whilst purely sexual goals were not (Berger-Merom et al., 2022).

> *“[Legitimate recovery goals] can be forming an intimate relationship but that does not necessarily have to be linked to sex I mean dating apps, going out for coffee and things like that are [legitimate since they are] still a step before sex.” (Berger-Merom et al., 2022)*.

### 1.3 Stigma against people with mental illness

Staff described stigma faced by individuals with mental illness from the staff themselves (Collins, 2001), the general public (Tiwana, McDonald & Völlm, 2016) or the patients’ families (Emery-Rhowbotham et al., 2024). Stigmatisation was perpetuated by the notion that service users should be protected and restricted from sex and dating, which might hinder rather than promote recovery.

> *“There’s so much stigma [about sex]. Stigma because they’re mentally ill. How can they want to have sex if they’re mentally ill? Also, they (the staff) see it as a symptom of mental illness—they’re hypersexual, so they must be protected from sex.” (Collins, 2021)*.

### 1.4 Considerations of the service users’ capacity for relationship building and maintenance

Staff felt that having conversations around intimacy may be challenging for service users (Southall & Combes, 2022), leading to the topic being omitted in therapeutic spaces entirely. Mental health nurses working at a long-term inpatient unit expressed concerns that patients may not have the mental capacity to consent to sexual relationships and this was reported as a reason for not supporting sexual intimacy between patients (Quinn & Happell, 2016).

> *“I think the assumption would be that for patients in acute, their mental state isn’t great and not stable and so they are in a state where they simply are unable to consent to be in a sexual relationship.” (Quinn & Happell, 2016)*.

In Uganda, there were reports of patients being administered family planning against their will, due to concerns of potential offsprings not receiving appropriate care (Tumwakire, Arnd & Gavamukulya, 2022).

### 1.5 Intimacy needs are not prioritised in mental health settings

Despite recognising the relevance and importance of intimacy in people’s lives, the need for romantic/intimate relationships was viewed as a peripheral issue in mental health settings. Staff reported that they felt unable to prioritise the topic due to limited time during consultations (Raisi et al., 2017), and tended to focus on other areas such as symptom management (Quinn, Happell & Browne, 2011).

> *“When someone comes in here often they’re … putting themselves or others at risk in some way and that tends to trump you know ‘how’s your relationship going how[‘s] your sex life going’ … and in a public system … you may only be seeing people in those times of crisis so you might not be building the relationship up to remember to ask … about all of those other factors.” (Urry, Chur-Hansen & Khaw, 2019)*.

## 2. Service provision at a personal level

Staff often reported a lack of knowledge and skills to offer support regarding intimacy needs, leading to feelings of discomfort or avoidance of the topic. As a result, they expressed a need for more training and guidance on how to have these discussions. Staff also sometimes reflected on their own experiences with intimacy and how this influenced their approach to the topic. This synthesised finding received a ConQual score of “low” and consisted of 43 findings which were divided into four categories.

### 2.1 Confidence of staff

Staff noted that they did not receive training on how to address intimacy and sexuality issues and expressed ambivalence regarding how to raise the topic in a safe manner (Forrester-Jones, Dixon & Jaynes, 2023). Staff also believed that they did not have expertise in managing and treating sexual problems, which led to them feeling ill-equipped to deliver support (Dubreucq, Lysaker & Dubreucq, 2023).

> *“When [sexual issues are] not talked about in training, what am I left with? Only myself, and what it triggers within me … so I react from my inner world … it’s not supposed to be like that … I don’t feel professional enough … because … I don’t have the tools.” (Berger-Merom et al., 2022)*.

However, staff from one study expressed feeling more confident and at ease with initiating conversations after more years of experience (Quinn, Happell & Browne, 2011).

### 2.2 Discomfort and avoidance with the topic of intimacy

The notion that intimacy was a taboo or overly personal subject was echoed by many of the staff (Forrester-Jones, Dixon & Jaynes, 2023).

> *“In our community, talking about sexual issues is very difficult for clinicians. How could a clinician educate patients regarding sexual issues when he/she does not feel comfortable in talking about these concerns?” (Raisi et al., 2017)*.

In particular, staff felt a heightened sense of discomfort to discuss intimacy with individuals of different genders, age, and religious or ethnic backgrounds from themselves, as these differences may be perceived as indicating that the individual would find discussions uncomfortable (Urry, Chur-Hansen & Khaw, 2019).

### 2.3 Self-reflections on experiences and beliefs relating to intimacy

Sometimes staff reflected on their personal beliefs, values and experiences related to sexuality and how this influenced their provision of care. For example, some staff reported reluctance to discuss homosexuality (Tiwana, McDonald & Völlm, 2016); other staff did not want to come across as stigmatising single status in service users (Emery-Rhowbotham et al., 2024). In Björn and Westman’s (2018) study, psychologists reflected on their experiences of navigating conversations with service users around sex, in which feelings such as frustration and satisfaction were acknowledged.

> *“I think it is important to reflect on your own judgements, because […] we’ve all got our own experiences, or thoughts around this and it does influence what you do.” (Urry et al., 2023)*

### 2.4 Having the right tools

Staff expressed a need for improved knowledge, training and access to an integrated service provision. Staff also reported wanting information on referral pathways for service users who require additional support for their intimacy needs, such as those struggling with their sexual identity (Urry et al., 2023). Staff felt training would reduce anxiety about broaching a “taboo” subject, raise confidence and decrease stigma among staff (Tennille et al., 2021).

> *“I think for people in the health system, [and] certainly mental health that are time-poor and having requirements to do everything else [… we should have] an online, course about sexual health and what it means and how to approach it with clients and ideas and tools to use and stuff.” (Urry et al., 2023)*

## 3. Fitting intimacy needs into the therapeutic context

This synthesised finding captured how staff conceptualised intimacy needs within the context of their therapeutic practices. Some staff believed that supporting people in finding relationships was not part of their role, while others felt that addressing intimacy needs would result in therapeutic benefits. Staff were also cognisant of the importance of building rapport and respecting the boundaries of a therapeutic relationship. A total of 42 findings divided into four categories made up this synthesised finding, which received a ConQual score of “low”.

### 3.1 The displacement of responsibility to raise conversations

Staff expressed uncertainty about whether intimacy needs fell within their scope of work, as they felt better equipped to handle mental health-related issues. Some believed that this responsibility should be addressed by others such as general doctors (Quinn, Happell & Browne, 2011) or staff with social roles such as peer support workers (White, Haddock & Varese, 2019). Moreover, service users were often expected to “set the agenda” by raising concerns around intimacy first (Björn & Westman, 2018).

> *“You stick to what you’re good at and you can maybe give ‘em a bit of advice but really you’re better off signposting to somebody who deals in that all day.” (White, Haddock & Varese, 2019)*.

### 3.2 The importance of nurturing and respecting the therapeutic relationship

Staff felt that building rapport with the service user laid the groundwork for productive discussions around intimacy and relationships. Being of similar age (White, Haddock & Varese, 2019), as well as taking time to establish trust were seen as facilitators (Quinn & Happell, 2012).

> *“Yes, I do (bring it up), but not always on the first contact. Usually I have some degree of rapport with them (consumers) first.” (Quinn, Happell & Browne, 2011)*.

However, staff noted that there were moral and ethical concerns which they felt restricted their ability to offer support around finding relationships (Emery-Rhowbotham et al., 2024). For instance, staff felt that they had the duty to actively discourage a service user from pursuing a romantic/intimate relationship, where this was perceived as hampering the person’s rehabilitative progress (Quinn & Happell, 2015a).

### 3.3 Addressing intimacy needs as part of recovery-oriented practice

Despite receiving limited training on how to offer support, staff believed that helping service users to build intimate connections facilitated recovery. Staff also suggested that having conversations on the topic would support service users in making more autonomous and informed choices about their sexual relationships (Dubreucq, Lysaker & Dubreucq, 2023). In addition, being in a relationship was often seen to be beneficial to the service users’ recovery trajectory, and may even encourage treatment adherence (Tumwakire, Arnd & Gavamukulya, 2022).

> *“It’s really important (to talk about sexuality). It’s a really essential part of people’s identity, their recovery, and their wholeness as human beings, to have the ability to love, be loved, and to express that intimacy with someone else, and to value themselves.” (Quinn & Happell, 2012)*.

### 3.4 Concerns about the impact on service users’ mental health

However, some staff believed that discussing intimacy would pose an emotional risk to patients’ vulnerable mental states (Forrester-Jones, Dixon & Jaynes, 2023). This was often rooted in their view that service user stability was the priority, rather than reflecting a resistance to addressing intimacy needs (Emery-Rhowbotham et al., 2024).

> *“The thing is, you know, this is a mental health facility and our goal is to provide care for their mental state, predominantly above the others, and if anything else that we feel might get in the way of harming that, then that’s what we try to minimise that as much as possible.” (Evans, Quinn & McKenna, 2019)*.

## 4. Service provision at an organisational level

Staff highlighted that mental health care tends to focus on risk management and safeguarding, which can make it harder for the intimacy needs of service users to be addressed. Staff expressed frustration with the lack of support to balance risk management with supporting people’s autonomy and advocated for better policies. This synthesised finding received a ConQual score of “low” and was made up of 46 findings grouped into five categories.

### 4.1 Restrictive clinical setting

In inpatient mental health settings, patients’ sexual activity was often put under great scrutiny. Some staff supported prohibitive “no-sex” policies (Evans, Holmes & Quinn, 2020), believing that hospitals were inappropriate places for sexual relations to occur (Tiwana, McDonald & Völlm, 2016). The high levels of surveillance and security were viewed as barriers to safe and healthy sexual expression among service users.

> *“Yes, I wonder myself how the patients manage it because if I’m in a room with two others, I find [masturbating] almost undignified […] But, as I said, this whole topic is rather under the radar right now. So no one is really thinking about it.” (Götzl, Büsselmann & Klein, 2023)*

### 4.2 Prioritising risk management and safeguarding

Staff viewed risk management as a key aspect of their roles. They were reluctant to support romantic/intimate relationships where they felt an individual was vulnerable to sexual exploitation (Quinn & Happell, 2015a).

> *“I think we’re probably more concerned about vulnerability issues – whether people are being exploited in any way – so they may get with a partner who may exploit them for money or other things.” (White, Haddock & Varese, 2019)*.

Intimacy needs were more likely to be perceived as relevant if there were safeguarding concerns. Staff were also mindful not to inadvertently encourage unwanted sexual advances from service users by raising the topic (Tumwakire, Arnd & Gavamukulya, 2022). Some however appeared to “turn a blind eye” to sexual activities that occurred outside the facility due to concerns of professional culpability in case something went wrong (Quinn & Happell, 2015b).

### 4.3 Counterproductive policies

Sometimes policies intended to safeguard staff were felt to do more harm than good, for example when staff were explicitly discouraged to discuss intimacy (Berger-Merom et al., 2022). Patients’ recovery goals relating to intimacy were often left unexplored as a result. Policies that were too rigid (Evans, Holmes & Quinn, 2020), too arbitrary (Evans, Quinn & McKenna, 2019), or too outdated were also seen to be counterproductive (Tiwana, McDonald & Völlm, 2016).

> *“The funny thing is when doing assessments, nowhere on the assessment sheet does it say anything about sexuality. There are no questions; there is nothing (asking) ‘Are you sexually active?’ Nothing … we avoid it like the plague (laughs)” (Quinn, Happell & Browne, 2011)*.

### 4.4 Lack of organisational support

Recognising the difficulty of addressing intimacy needs, staff stressed the importance of managerial support and supervision (Tennille et al., 2021). Yet, staff reported struggles with managing heavy workloads while receiving little guidance on how to integrate intimacy needs into routine practice (Tumwakire, Arnd & Gavamukulya, 2022).

> *“Everything you do has to be mostly approved or encouraged by them, so … without management support, it is not something that can be done.” (Emery-Rhowbotham et al., 2024)*.

### 4.5 The need for improved guidelines around intimacy needs

Staff believed that enacting change from an organisational level would positively impact service users. For example, providing a safe and private space in an inpatient setting for patients to cultivate intimate relations may deter them from taking sexual risks (Quinn & Happell, 2015b). Formal clarifications on staff’s clinical role in addressing intimacy was proposed as a beneficial idea, particularly if developed with staff input (Quinn & Happell, 2016).

> *“For the ones who are in relationships on campus it would be good because it would remove that element of sneaking around and doing it in places where human beings really shouldn’t be doing it … They could actually do it, do it comfortably and do it properly and get their needs met.” (Quinn & Happell, 2016)*.

## Discussion

### Main findings

The present review synthesised findings from 24 studies exploring mental health staff’s views and experiences with supporting service users’ needs for romantic/intimate relationships. The first synthesised finding discussed how staff acknowledged service users as sexual beings, an encouraging finding given that earlier studies reported some staff to consider service users as asexual (Tennille & Wright, 2013). However, our review found that staff often felt reluctant to talk about people’s needs and wishes for romantic/intimate relationships as part of their mental health care. This is concerning, given the common view among mental health service users that the opportunity to discuss romantic/intimate relationships with staff is appreciated and constructive (McCann et al., 2019). This difference in staff and service users’ perspectives was at least partially explained by staff struggling to balance their responsibility to safeguard service users whilst also providing them with support for this aspect of their lives. This synthesised finding also revealed how staff conceptualised sexual health and intimacy as distinct needs. Notably, sexual health was viewed under the framework of risk management while intimacy was more associated with recovery.

Staff’s lack of confidence in addressing intimacy needs was also a key finding. This has been explored in previous studies, and our findings were congruent with those of a similar review that investigated the perspectives of staff across healthcare professions (Dyer & das Nair, 2013). This may indicate the importance of the issue not just in mental health care, but across the healthcare sector. These results pose a strong imperative to improve training and develop guidelines for healthcare staff. There were some variations in what participants in our review considered as suitable training, with some viewing it as informational, while others desired practical tips on how to conduct conversations. Staff in the papers in our review were often reflexive about their personal values and experiences with sexuality, opening up on reasons why they might feel uncomfortable with the topic. A surprising finding was that cultural-based attitudes were only reflected upon in one included study (Raisi et al., 2017), despite previous research showing how culture influences our beliefs and treatments of intimacy (Marshall, 2008).

The third synthesised finding showed that staff were thoughtful about their roles and boundaries as mental health service providers, and described how intimacy needs might fit into their own approaches to support. However, there was evidence that staff often felt that addressing patients’ needs for intimacy was outside of their professional responsibility. This belief was shared by staff from primary care (Gott et al., 2004), suggesting that there may be a gap in addressing service users’ intimacy needs across different healthcare sectors. This is not surprising as human sexuality is reported to be absent from mental health professionals’ training (Wright, 2022). Given this, staff often adopted an exploratory approach to addressing intimacy based on standard therapeutic practices. Addressing intimacy needs was seen as no different than the pursuit of other therapeutic goals in terms of requiring the foundations of a good therapeutic alliance (Southall & Combes, 2022). This may partially explain why staff felt that service users should take the lead on initiating conversations on the topic (Pinto et al., 2012). Interestingly, research has suggested that staff-initiated conversations on sexuality may improve therapeutic relationships (Tennille, Solomon & Blank, 2010). Furthermore, our findings suggested that staff typically endorsed the principles of recovery-oriented care and identified intimacy as a facilitator to recovery, but some felt that in practice, intimacy was a delicate topic that should be reserved for when the patient’s condition has stabilised.

Finally, our review highlighted the growing research on the intimacy needs of service users over recent years, but also that little systemic change in service provision has occurred. Inpatient psychiatric facilities are typically governed by policies which prevent service users’ intimacy needs from being met, even where patients have capacity to make informed choices about sexual activity, and staff consistently reported a lack of support from their organisation to help them address this area. Recent studies involving NHS staff outlined the conflict between delivering compassionate care and the culture of care driven by proxy performance measurements (Bhui, 2016), such that staff have to prioritise interventions with more “clinical” than “social” outcomes (Emery-Rhowbotham et al., 2024). Additionally, staff expressed fatigue and frustration with expectations to deliver recovery-oriented care amidst staff shortages and large caseloads (Chatwiriyaphong et al., 2024). Our findings reflect on the extent to which mental health practice tends to focus on risk management rather than fostering the kind of therapeutic culture that empowers service users to cultivate relational resources (Tomlin, Bartlett & Völlm, 2018).

### Strengths and limitations

To our knowledge, this paper was the first to synthesise qualitative literature on mental health staff views on supporting service users’ needs for romantic/intimate relationships. The inclusion criteria were broad to capture the full spectrum of views, experiences and attitudes of mental health staff from diverse backgrounds and roles.

However, there are some limitations to this review. Although any study including staff who provide care to mental health service users was eligible for inclusion, the final sample consisted mostly of staff with clinical roles such as nurses, psychologists and psychiatrists. Our findings may therefore present a more medicalised narrative (Ahsan, 2020). Moreover, the included studies did not consistently supply information on staff’s ethnicity. Thus, important perspectives from ethnically diverse staff may not have been captured in this review. Only three studies reported on any PPI in the research process. This is an important omission as people with lived experience may highlight gaps in service provision through first-hand experience in navigating systems (Sunkel & Sartor, 2022).

In terms of methodological limitations, only four of the 24 papers we included were conducted in low-and-middle income countries (LMICs). Therefore, the context transferability of our findings is likely to be restricted. Although we defined intimacy needs clearly, many of the included studies examined sexuality, which is a related but distinct concept to intimacy. Furthermore, initial data synthesis was only carried out by one reviewer. While qualitative-evidence synthesis is inherently subjective, this could be seen as impacting the reliability of our findings. Finally, findings from all included studies were weighed equally regardless of their quality appraisal, although only one study was rated of moderate quality and none of low quality.

### Implications for policy and practice

The findings of this systematic review demonstrated that organisations need to build staff confidence through adequate training and support. Staff are often working in a policy vacuum, with little organisational guidance in whether, when and how they should discuss needs for romantic/intimate relationships with service users, or what support they might offer. Organisations should strive to create a culture of openness surrounding sexuality. Staff in our review felt that they had to dichotomise safety and sexuality, but evidence indicates that service users are better protected when they are empowered to speak up about sexual issues (Care Quality Commission, 2020). Service users should also be actively involved in their own care planning in regards to intimacy and relationships (Bee et al., 2015). Policies around staffs’ professional boundaries should be evaluated and reviewed to allow staff to understand what is expected of them and navigate the integration of sexuality in their conversations with service users. Apart from supporting staff in addressing intimacy needs, organisations have the responsibility to establish a working environment where staff feel safe and comfortable to carry out their work.

While trials for sexual health care training provided to mental health staff have shown promising results (Lu et al., 2024), sexuality should be routinely taught as part of staff training curriculum. Providers should be encouraged to bring up the topic of intimacy, and to reflect on their own beliefs and attitudes surrounding the subject (Wright and Pugnaire-Gros, 2010). Relevant staff development initiatives can be informed by campaigns created for other sectors, such as the “Supported Loving” toolkit which provides guidance for staff on how to support people with learning disabilities on issues surrounding sexuality and relationships (Bates, 2019). The impact of this campaign, evidenced by its growing network of almost 2000 members, underlines the potential value of similar resources and guidelines in mental health care.

Further, it is recommended that inpatient, community and social care sectors should collaborate to deliver holistic, patient-centred care, such as explicitly clarifying the responsibility of addressing intimacy needs and ensuring that specialist sexual health or relationship support can be accessed, in addition to supportive conversations with generalist mental health staff. Staff who specialised in addressing intimacy needs were not included in the review as it was not our aim, but previous research has recommended making couple counselling and psychosexual therapy more accessible to service users (McCann et al., 2019). This would require a concerted effort to delineate referral pathways.

### Future research

Sociocultural influences were noticeably absent in our review findings, and it would be useful to gauge staff views given the diversity in how different communities conceptualise intimacy and mental health care. As our review largely included clinical staff, there is a lack of qualitative studies examining the views and experiences of staff from mental health social care. The distinct recovery focus of mental health social care may introduce new perspectives on how to best deliver support surrounding intimacy. An update on McCann and colleagues’ (2019) systematic review on service users’ perspectives on what type of support they would like from staff would be helpful given the increase in research interest over the past five years.

More evaluation is needed for the development of new programmes and/or the adaptation of existing care models, such as the BETTER model for discussing sexuality in oncology settings (Mick et al., 2004), to mental health settings. The same goes for interventions that focus on service users’ romantic functioning (Cloutier et al., 2023), which can be a powerful addition to existing programs targeting the biological elements of sexuality such as safe sex, contraception and family planning.

## Conclusion

Our review has provided insight into mental health staff’s experiences and needs in relation to offering support for service users to engage in romantic/intimate relationships. Our review findings underscore personal, provider and organisational-level factors that influence the ongoing neglect of addressing service users’ intimacy needs in this domain in mental health settings. These have important implications for mental health care policy and boosting staff confidence in addressing intimacy through appropriate training and support.

## Data Availability

All data produced in the present work are contained in the manuscript.

## Acknowledgements

A huge thank you to my supervisors, Brynmor Lloyd-Evans, Helen Killaspy and Sharon Eager, for making this project possible. Thank you to Aisling Smith O’Connor for the support I received throughout the reviewing process. Thank you to Annabelle Olsson and Merle Schlief for their help screening and translating non-English papers.

## Data Availability

All data generated and analysed in this review are available in the manuscript and the appendices.

