## Appendices for "Exploring mental health staff’s views and experiences on supporting service users’ needs for romantic/intimate relationships: a qualitative systematic review"

**Appendix A**

***Search strategy***

MEDLINE

| # | Query | Results retrieved on 21/4/24 |
| --- | --- | --- |
| 1 | (Intima* OR romantic OR romance OR love OR loving OR sexual need* OR sexual relationship* OR emotional connection OR attraction).ti,ab,kf. | 136505 |
| 2 | (mental health OR mental illness* OR mental disorder* OR mental problem* OR behavioral health OR behavioural health).ti,ab,kf. | 323160 |
| 3 | exp mental disorders/ | 1485461 |
| 4 | exp mental health services/ | 107163 |
| 5 | (Staff OR worker* OR clinician* OR practitioner* OR provider* OR peer support* OR professional* OR nurse* OR doctor* OR care coordinator* OR personnel OR psychiatrist* OR psychologist* OR occupational therapist*).ti,ab,kf. | 1816426 |
| 6 | exp Qualitative research/ | 87077 |
| 7 | (view* OR perception* OR experience* OR attitude* OR understand* OR belief* OR interview* OR focus group* OR mixed method OR barrier* OR facilitator*).ti,ab,kf. | 4388335 |
| 8 | 2 OR 3 OR 4 | 1713981 |
| 9 | 6 OR 7 | 4396317 |
| 10 | 1 AND 5 AND 8 AND 9 | 1733 |

PsycINFO

| # | Query | Results retrieved on 21/4/24 |
| --- | --- | --- |
| 1 | (Intima* OR romantic OR romance OR love OR loving OR sexual need* OR sexual relationship* OR emotional connection OR attraction).ti,ab,hw. | 1111245 |
| 2 | (mental health OR mental illness* OR mental disorder* OR mental problem* OR behavioral health OR behavioural health).ti,ab,hw. | 392951 |
| 3 | exp mental disorders/ | 1110629 |
| 4 | exp mental health services/ | 59790 |
| 5 | (Staff OR worker* OR clinician* OR practitioner* OR provider* OR peer support* OR professional* OR nurse* OR doctor* OR care coordinator* OR personnel OR psychiatrist* OR psychologist* OR occupational therapist*).ti,ab,hw. | 1000000 |
| 6 | exp Qualitative methods/ | 22073 |
| 7 | (view* OR perception* OR experience* OR attitude* OR understand* OR belief* OR interview* OR focus group* OR mixed method OR barrier* OR facilitator*).ti,ab,hw. | 2379023 |
| 8 | 2 OR 3 OR 4 | 1306559 |
| 9 | 6 OR 7 | 2382840 |
| 10 | 1 AND 5 AND 8 AND 9 | 3032 |

Web of Science

| # | Query | Results retrieved on 22/4/24 |
| --- | --- | --- |
| 1 | (Intima* OR romantic OR romance OR love OR loving OR “sexual need*” OR “sexual relationship*” OR “emotional connection” OR attraction).ts. | 421978 |
| 2 | (“mental health” OR “mental illness*” OR “mental disorder*” OR “mental problem*” OR “behavioral health” OR “behavioural health”).ts. | 466670 |
| 3 | (Staff OR worker* OR clinician* OR practitioner* OR provider* OR “peer support*” OR professional* OR nurse* OR doctor* OR “care coordinator*” OR personnel OR psychiatrist* OR psychologist* OR “occupational therapist*”).ts. | 2624534 |
| 4 | (qualitative OR view* OR perception* OR experience* OR attitude* OR understand* OR belief* OR interview* OR “focus group*” OR “mixed method” OR barrier* OR facilitator*).ts. | 9593405 |
| 5 | 1 AND 2 AND 3 AND 4 | 2398 |

CINAHL

| # | Query | Results retrieved on 22/4/24 |
| --- | --- | --- |
| 1 | (Intima* OR romantic OR romance OR love OR loving OR “sexual need*” OR “sexual relationship*” OR “emotional connection” OR attraction) | 50534 |
| 2 | (“mental health” OR “mental illness*” OR “mental disorder*” OR “mental problem*” OR “behavioral health” OR “behavioural health”) | 252095 |
| 3 | (“mental disorders”).mh. | 68815 |
| 4 | (“mental health services”).mh. | 38043 |
| 5 | (Staff OR worker* OR clinician* OR practitioner* OR provider* OR “peer support*” OR professional* OR nurse* OR doctor* OR “care coordinator*” OR personnel OR psychiatrist* OR psychologist* OR “occupational therapist*”) | 1552765 |
| 6 | (“Qualitative studies”).mh. | 153599 |
| 7 | (view* OR perception* OR experience* OR attitude* OR understand* OR belief* OR interview* OR “focus group*” OR “mixed method” OR barrier* OR facilitator*) | 1559837 |
| 8 | 2 OR 3 OR 4 | 252095 |
| 9 | 6 OR 7 | 1574669 |
| 10 | 1 AND 5 AND 8 AND 9 | 964 |

**Appendix B**

***Quality appraisal tool***

1.	Is there congruity between the stated philosophical perspective and the research methodology?

2.	Is there congruity between the research methodology and the research question or objectives?

3.	Is there congruity between the research methodology and the methods used to collect data?

4.	Is there congruity between the research methodology and the representation and analysis of data?

5.	Is there congruity between the research methodology and the interpretation of results?

6.	Is there a statement locating the researcher culturally or theoretically?

7.	Is the influence of the researcher on the research, and vice- versa, addressed?

8.	Are participants, and their voices, adequately represented?

9.	Is the research ethical according to current criteria or, for recent studies, and is there evidence of ethical approval by an appropriate body?

10.	Do the conclusions drawn in the research report flow from the analysis, or interpretation, of the data?

**Appendix C**

***Extracted findings and illustrations***

(U) = unequivocal level of plausibility (finding is accompanied by a quote that is beyond reasonable doubt)

(E) = equivocal level of plausibility (finding is open to challenge)

(Un) = findings are not supported by the data

|  | **Berger-Merom, R., Zisman-Ilani, Y., Jones, N., & Roe, D. (2022). Addressing sexuality and intimate relations in community mental health services for people with serious mental illness: A qualitative study of mental health practitioners’ experiences. *Psychiatric Rehabilitation Journal*, *45*(2), 170.** | |
| --- | --- | --- |
|  | Finding | Illustration |
|  | Lack of training to discuss sexuality and intimacy issues (U) | *When [sexual issues are] not talked about in training, what am I left with? Only myself, and what it triggers within me … so I react from my inner world … it’s not supposed to be like that … I don’t feel professional enough … because … I don’t have the tools.* |
|  | Uncertainty about whether intimacy falls within their scope of work (U) | *Being a psychiatric rehabilitation practitioner, am I in a position to discuss an issue that I don’t know how it will develop? Is it within my realm of expertise? And if there’s sexual trauma … who am I to deal with it? Is it still part of rehabilitation or should it be part of psychotherapy?* |
|  | Discomfort with discussing sex (U) | *A client … told me he had sex for the first time, [and] described it in detail. It was too much for me, I [could not] bring it to [the] supervision meeting ... I avoided it, so I chose not to bring this to supervision.* |
|  | Desire for romantic relationships is seen as a legitimate recovery-related goal, while purely sexual needs are not (U) | *[In my opinion, legitimate recovery goals] can be forming an intimate relationship but that does not necessarily have to be linked to sex. ... I mean dating apps, going out for coffee and things like that are [legitimate since they are] still a step before sex ... they did not get there yet.*  *[In my view] [a] consumers’ main goal cannot be a [purely] sexual one. [A main recovery goal] can be related to improved self-esteem, losing weight or going to the gym, but it cannot be paid sex [i.e., prostitution].* |
|  | Sexual decisions that affect others are seen differently (U) | *Ethical issues if ... someone wants to get pregnant.The question is ... is she able to raise a child? Our approach to supported housing is that we are ... part of the fabric of their lives, helping them make decisions, advising, but [we] don’t decide for them. But this ... having a child ... raises major dilemmas [about] how much we get involved.* |
|  | Program-level policies are counterproductive to navigating client goals to seuxality (U) | *There’s a rule here that … we are not allowed to talk with tenants too much about sexuality as paraprofessional staff … I think it’s for the best … it protects us … we’re with them all the time, including night times … A tenant can complain about sexual harassment and say … “The care worker sat with me until two in the morning and talked to me about sex … I was harassed” … Go prove it didn’t happen.* |
|  | Education programs focus on sexual health instead of intimate relationships (U) | *In hostels [residential programs] clients have sexual education [sessions] … once every 6 months … we talk about condoms … [HIV/ AIDS] … how to have safe sex … less on how to approach and achieve sexual and intimate relations.* |
|  | Providers are exposed to awkward situations relating to patient's sexuality (U) | *A 65-year-old tenant .. always asks [for my help] to search porn videos on the computer for him … so I would do that and walk away. Two weeks ago [I] needed to [help him in his] cleaning duties [in the residential program]. I knocked on the door, opened it and saw him [in a middle of a sexual act] with a sex magazine on the table and [his] pants down … I said, “Excuse me, but you promised to do your chores, you’re avoiding it all day.” He looked at me and said, “Can’t you see I’m busy?” … [Although] he and I have a pretty open interaction level, I [ignored the situation and asked] him [to] “just finish whatever you [do] and come do your chores; I’ve been nagging you about it all day already.”* |
|  | **Björn, T., & Westman, S. (2018). Distribution of Power in a Jumble of Emotions: Clinical Psychologists’ Experiences of Talking with Patients About Sex and Sexuality.** | |
|  | Staff felt a professional responsibility to guide and protect the patient (E) | *"Yes, but you think that, I think what actually makes me feel reluctance is that I don't want the patient to, that the patient shouldn't react, "what the hell, I don't have anyone to sleep with, why are you asking! And that it might be a bit like… like a rub-it-in question but also like… I'm missing a body part and well, how does it feel to have it? A bit like… but I still think that even though I have to think actively, it's still good that I ask because, for some, it's still a problem with desire on its own and for some, it's still a great source of joy and quality of life to have it."* |
|  | The experience of conversations often revolved around trying to understand the patient (U) | *" when I was little like that curiosity. So, to try like, let’s see if we can find some bad analogy here ... when I was a kid, I liked to unscrew things to see how they worked ... I feel like it’s the same in the patient room, so unscrewing bits here and there to see how this person works. So pretty neutral, but curious.* |
|  | Staff viewed themselves as leaders in the therapeutic process (Un) | *"I think that on the one hand it's good to normalize that it's normal to talk about sex, but it also feels like it's good to normalize that you're not feeling so well right now, it might not be so strange that you're not having sex with your partner right now, because that's also normal. So I mean, you don't want to put any more pressure on yourself - everyone has to have sex, because if you're super stressed or generally feeling a little bad, maybe that's not what's at the top of the agenda, you might still have a loving relationship with someone and get as much support as possible."* |
|  | Topics surrounding sex and sexuality were engaging and important to work with (U) | *Yeah ... I leaned forward, I maybe felt like, how should I say, like an ... arousal, although not sexual arousal, more like this is engaging, like an upper, became alert in the head, to feel like your blood pressure goes up a bit, like this is relevant, we’ve got something here, so, wow, this seems to really hit a nerve, we could make a thing out of this, it feels important and I feel sharper or what should I say...* |
|  | Patients who were open to broaching difficult subjects were seen as facilitating the work of the staff (U) | *"… no but that was right, they were two quite different people, but still you felt that they were an open person who can talk about anything, who doesn't shy away from difficult topics and who brings up difficult things and that it's natural that it feels good. So that it's nice."* |
|  | Staff adopted a listening approach and allowed patients to take control of conversations (U) | *"What's important to me in a situation like that is not to change the subject, not to try to... like turn it into something else, like, it's clear that, that, what it's like for a person who seeks their partner for sex and gets rejected... that... that can rightly be associated with a lot of other things like and... in this relationship and about what it's like to meet people and have your needs met or not met... but, to, to just stop somewhere at, at exactly what she says, she talks about sex, then we talk about sex and then we can, then we can take this later... Similes, or like comparisons or like, that'll have to wait."* |
|  | Conversations should take place on patients' terms and their own language (U) | *"And I don't attach importance to it unless they do. If it feels important in some way in their identity building or if it affects relationships or, well, then we can talk about it. But if the person themselves is, well, this is how I am but I need help with my compulsion and it doesn't affect the sexual aspect at all, well, then we'll sort it out."* |
|  | There was a fear of asking questions which were too private or uncomfortable (U) | *"… again that the patient might feel offended. Ah, but that the alliance is somehow going to be broken because of some kind of transgression, I think. It's those kinds of concerns that are holding me back. Oh, I think that if the patient wants to talk about it, they'll bring it up."* |
|  | Staff felt more comfortable with having a standardised way of asking questions regarding sex and sexuality (U) | *"I have, I'm the kind of person who, when it comes to sensitive questions, blames it on having to ask everyone the same thing. So I have a printed out medical history sheet and so, now I will ask about important things in life like, everyday things that affect many people, how is food, how is sleep, how is sexuality and sex and if they bring up that they have a partner then I say it is a good relationship and then I ask how the sexual thing works..."* |
|  | Staff felt a need to set boundaries in conversations with patients (U) | *"… that's what I didn't want to do, or that's probably a bit more that I, ah, myself, ah, distanced myself a bit in some way … And I don't know how but, I absolutely wouldn't, with some patients I can say something a little personal, you reveal something a little to get deeper, further, but I wouldn't do that with him."* |
|  | The topic of sexuality was perceived to be difficult and challenging (U) | *"So then we talked a lot about it, and I thought it was a little strange or hard to deal with because, or I don't know... asexuality is difficult, for me, or I understand that there are people who don't want to have sex, that it's something you don't want to do... but it's, it's very easy, I experienced at the beginning of the case that I just interpreted it as this being an avoidance behavior."* |
|  | Staff expressed powerlessness as they didn't know how the patient can be best-helped (E) | *And so I gave tips on where you can, can seek further help just to say that there is family counseling, it is here, you can go individually, you can search online, so like that, just brainstorm. Encourage continued learning. Something like that. Or also, also encouragement to discuss with your doctor if you are experiencing difficulties too and that there are those who are more specialized that you can seek. Seek more information from.* |
|  | Staff engaged in self-reflection about different situations they encountered (U) | *"So I don't think it turned out bad. So that, it doesn't feel like, I don't carry it with me like some kind of, stone in the shoe feeling, oh what a bad thing it turned out to be... eh, but... but of course there can be a wonder if it could have been, like more, although I don't know! So. So that it is, therefore it is like... well a little neither or in some way. But I, I think that... well, it's also in a context so that I thought it turned out, I think it was good."* |
|  | Staff were sometimes caught off guard when engaging in conversations around sex and sexuality (U) | *"I might … be a little surprised by everything he tells me and I certainly believe it's true but […] but what is this really like, what kind of sexuality can drive so very hard, both on their part and on his part and what is the driving force behind it?"* |
|  | Concerns arose when the professional therapeutic relationship shifted to something more private (U) | *Because I didn't feel comfortable with him, in a way, and it was probably just how I imagined it, but I thought of myself as an object or that I became one of all these women that he looked at or like that I felt a little like... ah, but I thought I wonder if he now looks at me, ah, in that more objectifying way that he did with so many others.* |
|  | Conversations around sexuality felt natural and like any other subject (U) | *"I would have thought it would have felt harder because we had a long history of not talking about sex, that it suddenly becomes a big topic that wasn't even on the map before. It was like nothing we mentioned other than in passing like this sometime in the beginning, no but it's no problem it works or like this nothing special, until it became quite in-depth, it was quite detailed. It was a big step but that.. I also think that we had something like this that we could talk quite openly or very openly, so used to each other in a way so it wasn't that strange […] it felt natural"* |
|  | Staff reported a range of emotional experiences based on their experience of control over the conversation (U) | *"Towards the end it probably became a bit like this, when he became a bit more private with me, partly I got irritated, but also a bit scared, because he was a bit unpleasant, and the last conversation he didn't want to leave when I said we were done, because then I had said we would go on for an hour and we were done after 45 minutes […] And then we came to the conclusion that it was all a lie, he had never met these women … So then it also became like this, what was that, I still don't know what it was … But then it also becomes very doubly, I can get irritated and frustrated that, that's what the hell you're doing!"* |
|  | **Collins, P. Y. (2001). Dual taboos: sexuality and women with severe mental illness in South Africa. Perceptions of mental health care providers. *AIDS and Behavior*, *5*, 151-161.** | |
|  | Family planning is encouraged (U) | *[Family planning] is something I promote. I don’t have any problem with the individual having a child or family. I don’t see my opinions for people with MI as separate form other people. I also think people with SMI who can’t sustain periods of mental health should take measures not to have kids. A lot of people are off meds when they’re pregnant. As soon as the child is born they need to be back in the hospital. You need more assistance. It requires more resources. I suppose there’s the whole genetic argument—one must be honest. Yes, there’s a chance your child may develop a mental illness, but there’s a chance my child will, too.* |
|  | Lack of confidence in addressing needs for intimacy (U) | *One of the questions I ask is what to do about our patients’ need for intimacy. We have a fear of addressing this. The fear is to what extent can psychiatric patients give consent for sexual activity. But a psychiatric patient is a human and they will have needs. How do we choose their partners? Especially in chronic institutions. ...* |
|  | Stigma against intimacy needs in patients with mental illness (U) | *There’s so much stigma [about sex]. Stigma because they’re mentally ill. How can they want to have sex if they’re mentally ill? Also, they (the staff) see it as a symptom of mental illness—they’re hypersexual, so they must be protected from sex. Not all patients who want to have sex are hypersexual. There’s also a moral judgement in it. Sex is something exclusive and these people are mentally ill—they’re like animals.* |
|  | **Dubreucq, M., Lysaker, P. H., & Dubreucq, J. (2023). A qualitative exploration of stakeholders’ perspectives on the experiences, challenges, and needs of persons with serious mental illness as they consider finding a partner or becoming parent. *Frontiers in Psychiatry*, *13*, 1066309.** | |
|  | Importance of recovery-oriented practice (U) | *Peer-worker1 (PW1): “For me, intimate relationships and parenting, it’s also being able to make a free, informed choice (…) To choose by oneself, telling: “for me, this will be a good idea, this will be a rewarding experience.” (…) To make an informed choice, you need to have accessible information”* |
|  | Stigma against service users' ability to form intimate relationships (U) | *Nurse2: “There is some kind of background, (…) well, eugenism, to not discuss that because he would not be able to a good parent”* |
|  | Power differentials between provider and patient were a barrier (E) | *SW1: “We care for people who live under the medical power for years.”* |
|  | Patients may find it difficult to disclose mental illness to intimate partners (U) | *Psycho1: “Disclosure: how to tell it, when, to what extent, at which point of the relationship.”* |
|  | Feelings of discomfort, loneliness or resourcelessness related to a perceived inability to provide adequate support (U) | *PW1: “I think we’re a bit left alone on that (…). I mean, when some people come to us with some serious issues such as custody of children, foster care (…) What do we do?* |
|  | Need for improved knowledge, training, and access to an integrated service provision (U) | *Psychiatrist 1: “maybe a space for providers (…) we don’t have the entire network in mind”* |
|  | **Emery-Rhowbotham, A. A. I., Killaspy, H., Eager, S., & Lloyd-Evans, B. (2024). Finding a Relationship Conversations Between Mental Health and Social Care Staff, and Service Users. *medRxiv*, 2024-05.** | |
|  | Having conversations encourages recovery (U) | *"I feel that romantic relationships and intimacy are a human need and have a huge impact on mental health" [ppt. 36]* |
|  | Support regarding intimate relationships is relevant to service users (U) | *"I believe this is important. Because many service users can struggle to form relationships in general, and having some support regarding intimate relationships would be useful for these users" [ppt. 58]* |
|  | Finding a relationship is beyond staff's scope of work (U) | *"I think it's only our work role if its [mental health] related" [ppt. 29]* |
|  | It would be unethical to help service users find a relationship (U) | *" I feel like it would be unethical to help them find a relationship" [ppt. 3]* |
|  | Relationship seeking support is perceived as inappropriate (U) | *"Feeling it is inappropriate in my work role." [ppt. 2]* |
|  | Doubts about service users' ability to communicate their needs (U) | *"The person who we support being able to properly communicate their desire for a relationship even if it is something they might want" [ppt. 13]* |
|  | Lack of training on how to discuss intimacy (U) | *"I have experienced no training or discussions around this therefore naturally you think it might be out of the scope of your professional boundaries..." [ppt. 25]* |
|  | Lack of support (U) | *"everything you do has to be mostly approved or encouraged by them, so … without management support, it is not something that can be done” [ppt.5]* |
|  | Lack of policy (U) | *"Relationship goals not being part of routine assessment" [ppt. 46]* |
|  | Lack of time and resources (U) | *“there is barely enough time to do the core aspects of my job, so … there is very unlikely to be resource for this” [ppt. 43]* |
|  | Belief that relationships are inappropriate for the service user (U) | *"the individual may not be socially capable of a relationship e.g. if they display traits of aggression" [ppt. 5]* |
|  | Service users must first be stable (U) | *"Sometimes people, at least in more acute services, might benefit from more stability (e.g. of mood, of routine, being able to go out) if they are then to find a good relationship." [ppt. 51]* |
|  | Societal stigma against people with mental illness dating (U) | *"the general perception of the public about people with mental illness dating" [ppt. 33]* |
|  | Families being overprotective (U) | *"If the family are heavily involved, they can often be overprotective and not want their child to be dating" [ppt. 5]* |
|  | **Evans, A. M., Holmes, D., & Quinn, C. (2020). Madness, sex, and risk: A poststructural analysis. *Nursing Inquiry*, *27*(4), e12359.** | |
|  | Sexual activity was strictly policed by clinicians and in accordance with the ‘policy’ (U) | *…There is a very strict policy here that all social en-gagement between males and females has to take place in common areas… (Danielle).* |
|  | Heterosexual normativity of 'no-sex' policies (Un) | Not Supported. |
|  | Policy as a way to secure harmonious therapeutic environment (E) | *…There is a very strict policy here that all social en-gagement between males and females has to take place in common areas…[She] had her partner over and they were sitting in the lounge room watching television in a bean bag with a blanket over them and were told that this was wrong and that they couldn't do this… Because he was in her unit, and it was sug-gested that they go up to the rec room… and bedroom doors are shut. And this is respected…. But the main aim is not to have people doing that in the lounge room… (Danielle).* |
|  | Restricting intimate relations on-site endangers the safety of patients (U) | *…This really concerns me because like where do you go? If the person they are seeing lives in the same sort of environment as this or they may live with their parents and don't want to have sex in their parent's house or they are not allowed to or you know, there are a whole lot of, I mean. Yep that is pretty much what I am saying. They could be going off to parks or public toilets because they are in a relationship and they are wanting to have sex and it's just kind of…(Quentin)* |
|  | **Evans, A. M., Quinn, C., & McKenna, B. (2019). The governance of sexuality in a Recovery‐oriented mental health service: Psychosis, consumers and clinical approaches. *Journal of Psychiatric and Mental Health Nursing*, *27*(2), 194-202.** | |
|  | Service users' sexual activity is highly scrutinised (U) | *…I mean the staff have keys to clients’ units and bed -rooms and staff are often doing obs and wanting to know where clients are and what they are up to and it is very much like a constant check‐up from the point of view of the clients…[A.2.3]* |
|  | Viewing service users via a psychiatric lens (U) | *…. The thing is, you know, this is a mental health facility and our goal is to provide care for their mental state, predominantly above the others, and if anything else that we feel might get in the way of harming that, then that’s what ‐ we try to minimise that as much as possible…[G.3.1]* |
|  | Vague and arbitrary guidelines on 'inappropriate sexual behaviours' (E) | *…And yeah, I kind of was lucky enough to have a client who, like, part of his illness, like when he relapsed, is that he become sexually inappropriate so I could make it a really clear goal…[J.1.3]* |
|  | Service users' sexual behaviours are no different than ours (U) | *…I’ve seen some things documented that don’t necessary; I mean they are normal things that people would do. You know? I mean nothing comes to mind right now but I remember thinking ‘I do things like that’ and um and I feel that, that would be something that they could potentially write up in their notes. Like is it really relevant I guess…[P.6.1]* |
|  | **Forrester-Jones, R., Dixon, J., & Jaynes, B. (2023). Exploring romantic need as part of mental health social care practice. *Disability & Society*, 1-23.** | |
|  | Romantic relationship needs are only addressed if there are safeguarding concerns (E) | *Phillipa: ‘And then we get into “so what kind of skills would you need to develop a relationship?” “And what kind of situations would you put yourself in to meet someone?”’ (FG6)* |
|  | Lack of knowledge in how to deliver support (U) | *Toni: ‘Yeah, I would too. I wouldn’t be sure like how to deliver it within boundaries because it can get quite personal. So, I’d wonder how to approach that in the safest way. I think it’s really important to have relationships I think’ (FG6).* |
|  | Conversations may cause patients to deteriorate (U) | *Phillipa: ‘[…] So it feels like when you’re working it’s, when you look after a plant it’s almost like you’ve got to the stage where the shoot is really tiny and delicate and it’s like kind of, for me the relationship is like a tsunami could come through very fast and anything that has been brought up can deteriorate’ (FG6).* |
|  | Avoidance of and discomfort with discussing sex with service users (U) | *Phillipa: ‘I think with me there’s only so much I want to know about other people I support. I used to support an elderly woman who was too open about her hygiene habits than I actually wanted to know and felt I needed to be involved with. Like it’s a little bit like that with sexuality also with that there’s some things that I’m thinking, “do I really want to be involved with that?” Um so I think there’s something about personal comfort zone’ (FG6).* |
|  | **Götzl, C., Büsselmann, M., Klein, V., Streb, J., & Dudeck, M. (2023). Sexuality in Forensic Psychiatry. Results of a Qualitative Study on Professionals' Perspectives and Recommendations for Clinical Practice. *Psychiatrische Praxis*.** | |
|  | There were no clear guidelines regarding how to systematically address sexuality in everyday hospital life (U) | *"How is, in your opinion, the sexuality of the patients considered in your institution? Is it considered at all?" (Interviewer, MB)   “Yes, it is. But in a way that is still too unstructured […]. As I said, there is one document that mentions ‘sexual history/anamnesis’. But it’s rather a matter of discretion/personal judgement what exactly should and shouldn’t be included there.” (GZ_26)* |
|  | Sexuality was only taken into account if there were concerns such as the risk of pregnancy or abuse (U) | *"No, [sexuality; anamnesis CG] is not taken into account. The 'collateral damages' have to be taken into account if they [...] come back pregnant, if a pop-up tent used as a sexual refuge (hiding place for sexual activities) is found somewhere along a passage/doorway or a blanket or some condoms or, who knows, the relaxation of a sentence/restrictions is being abused [...]. For me, these are 'collateral damages' that arise from not paying attention/not taking sexuality into account." (TK_43)* |
|  | Patients often benefit from having sexuality explicitly discussed (E) | *“Well, actually the goal is for the patients to learn not only how to deal responsibly with their bodies and addictive substances, but also with their sexuality. And there […] it is important to learn to set boundaries appropriately and to get a feeling for their sexuality and […] to be able to defend themselves against prostitution.” (TK_46)* |
|  | Therapeutic processes should respond to the individual need of patients (U) | *"Of course, I can imagine many cases where there are big differences in terms of the experience of patients under section 63 compared to section 64. [...] You don't have to tar everyone with the same brush [...] even within section 64. Rather, there too, you should always leave the option open of being able to react or respond to the individual situation. (TK_41)* |
|  | Satisfaction of sexual needs was perceived to be harder for patients with longer lengths of stay (U) | *"I think that the topic, in my opinion, is much more present among the '63s, because the '64s, they know that they are getting out in two years [...], some of them, well the majority, actually are in a relationship outside (of the clinic), have a wife, some even have family [...], are meeting women and start a new partnership, yes. I think this suppression of sexuality is not as pronounced as it is among the '63s. They don't really have the chance to get to know someone or, um, yes, or start a new relationship and most of the time, um, the contact to the outside world gets lost over the years." (GZ_37)* |
|  | Patients experience a lack of privacy, sometimes due to overcrowding (U) | *"Yes, I wonder myself how the patients manage it because if I'm in a room with two others, I find it almost undignified [...], so if I don't already have a partner, [masturbation; note CG] is rather sparse here, yes. But, as I said, this whole topic is rather under the radar right now. So no one is really thinking about it." (TK_49)* |
|  | Visitation areas do not explicitly enable or promote intimacy (U) | *"And I would actually like a clear line. So either it is not allowed, then it is not possible, or it is allowed, then I have to say, hm, then please use the left one and somehow there are wet wipes to clean up afterward, it is such a shady gray area, too vague and I don't like that." (GZ_34)* |
|  | Parenthood was viewed as a controversial topic (U) | *"There [...] opinions diverged a bit. It was always a controversial topic, where everyone said, a child does not belong in forensic psychiatry [...] the children, [...] they then basically grow up among criminals." (TK_33)* |
|  | Prostitution was predominantly viewed with skepticism (U) | *"I have to honestly say, the first impulse is somehow skeptical because it would also have to do with promoting prostitution at the same time. As I said, meeting room, if it exists and if it could be made possible for any partner to come, no matter who it is, to live out sexuality, yes. Promoting prostitution somehow, I find difficult. I would have to inform myself and get clarification legally first [...] (TK_45)* |
|  | Staff were careful to maintain a professional distance from patients (U) | *"The first trap is that we live with the patients. A nurse works in a three-shift system. That means he sees the patients more often than his partner at home. And that is at least one year up to twenty years. That means there is a closeness that you sometimes don't even have in your own partnership." (GZ_33)* |
|  | Staff were aware of procedures to follow in the event of known sexual assault against staff (Un) | *N/A* |
|  | A careful approach to sexuality is needed for sex offenders (U) | *“There is always this difficulty that sex offenders always look for a weaker link, sexually speaking. […] And I'm a bit sceptical about how they deal with it [...] when you say, OK, sexuality is allowed here, is that a free pass, I'll touch everyone now [...], so really showing them the boundaries and rules." (GZ_37)* |
|  | **Hughes, E., Edmondson, A. J., Onyekwe, I., Quinn, C., & Nolan, F. (2017). Identifying and addressing sexual health in serious mental illness: views of mental health staff working in two NHS organisations in England. *International Journal of Mental Health Nursing*.** | |
|  | It was difficult to differentiate between safeguarding issues and acceptable lifestyle choices (U) | *“There's something in our training, in our experience, our social expectations. (Pause) and I think when people bring dilemmas about relationships and their sex lives to team meetings, I think sometimes they're the most difficult discussions, cos it's, you know, it, it might, on the face of it, be a very bad relationship, but is it really for us to say? It's very awkward. It's, it's a dilemma, I know, on inpatient units, but perhaps even more so in the community where, you know, the duty of care is less apparent and more distant, but actually still exists.” Participant 6, group 1, female* |
|  | Service users who were perceived as being more high risk were more likely to receive sexual health care (U) | *“It’s a real bad bias, and it’s a stereotype, but if you had a, a young gay man you’d probably be more alive to it [sexual health care] than if you had a young person, another young person” participant, 2, group 4, female* |
|  | Staff were aware of sexual health needs of service users (U) | *“Issues of either disinhibition due to mania or impulsivity and maladaptive or self-defeating coping behaviours resulting in unprotected sex, STIs, unplanned pregnancy, particularly significant in people who are given mood stabilisers” Participant 1, group 2, male* |
|  | Addressing sexual health was less of a priority in clinical practice (U) | *““it’s in the top five, for me, along with like dentistry, smoking, obesity, I don’t know, sedentary lifestyles and sexual health, the only thing that keeps it from, I think it’s probably at the bottom end of that top five only because people are generally very isolated, a lot of our patients are very isolated and have very poor quality sexual lives. Working within the community, you know, it’s, a very common recurring complaint is loneliness, lack of, you know, intimate relationships in general, which, which sexual life is, is a component of …. so from a service side of things, you know, that’s, that’s not as concerning to us as sort of an unfulfilled thing, so it’s only cos it’s not dangerous” participant 1, group 3, male* |
|  | Sexual health provision in mental health services was limited (E) | *“No it certainly isn’t something I ask routinely, It probably is something that I should (pause)…because even the less toxic medications that we prescribe, it’s probably worth knowing whether somebody is at risk or planning or thinking about the possibility of falling pregnant.” Participant 1, group 2, male* |
|  | Staff felt a lack of knowledge and confidence in broaching the subject (U) | *“this is outside normal practice, so, for both parties, so from the staff member’s point of view, if this person’s not used to being asked this, or it isn’t something that is routinely asked and I’m, you know, no-one wants to go out on a limb with anything or no-one wants to sort of be doing something that deviates from the norm, so if it’s not routine, if it doesn’t form part of routine questioning to be questioning somebody about an intimate part of their life, you know, out, outside of the framework of what I’m supposed to do during my assessment makes you feel uncomfortable, like, you know, you’re untested waters, you know” Participant 1, group 3, male* |
|  | Staff were concerned about causing distress to the service user and damaging the therapeutic alliance (U) | *“perhaps, you know, females and, you know, that’s something that perhaps, if, in case they have experienced, you know, trauma and abuse when they was younger or, and it was a male, but I suppose equally men suffers from abuse as well; so maybe it is a gender thing and it’s something that I, I’m just aware of, you know” participant 3, group 3, male* |
|  | Making sexual health a part of routine enquiry would support conversations around the topic (E) | *“We give health information and promotion advice on a whole load of things don’t we? But we don’t really on sexual health; and in a way we kind of, I don’t know, need permission to do that” Participant 1, group 4, female* |
|  | It was important for mental health professionals to address sexual health as part of their provision of holistic care (U) | *"Actually it is something that we need to address, because it does affect a lot of other sort of things about the person’s care. Remember we talked about, initially, safety, the young person’s safety, their physical health and then relationships, and then sort of their future, the children. So yeah, I think you, you know, it’s really important for us to address it, cos it’s not just sexual health, it’s sexual and other things around that person” Participant 4, group 4, female* |
|  | There was a need for training for all staff groups (U) | *“The stuff we cover now is diabetes, you know, bit of coronary heart disease if we, you know, we don’t, there’s no sexual health content actually in any of the post-qualification stuff that we do” participant 1, group 3, male* |
|  | **Lindskog, A., Lindroth, M., Holmgren, K., & Gunnarsson, A. B. (2024). Balancing on a slack line–Staffs’ experiences of talking about sexuality and sexual health with patients cared for in forensic psychiatry in Sweden. *Frontiers in Psychiatry*, *15*, 1450377.** | |
|  | Conversations around sexuality are seen as risk reducing in terms of both compliance to treatment and potential relapse (U) | *This is something we have to work with, it’s like our responsibility to help in some way. But if you have more of this holistic view of the human being, then it’s part of the whole, and then perhaps more is included for some individuals and less for others. And then it must be something you have to take responsibility for (P18).* |
|  | The mission to protect the society is often prioritized higher than patients’ individual needs and rights for care (U) | *Society, what we do to protect society is seen to be so very important. Sometimes, I think we, we sometimes abandon our patients, thanks to this protection that we somehow assume we are responsible for. And then it becomes more monitoring than nursing (P15).* |
|  | It is important to normalise sexuality (U) | *Address the problem with side-effects and the like, you may need to bring it up a little, a little bit and then pick it up again. You often have to split it up a bit into different small parts, but also to actually address that, that it is possible to have a normal sexuality, that it doesn’t have to be a problem, even if you’re locked up in forensic psychiatry (P3).* |
|  | Staff had to be responsive to conversations regarding reproductive health and relationships (Un) | *How I’m ever going to be able to find someone when I’m in here in a secure unit, or How will I ever be able to find someone when I’ve been in a secure unit, no one will want to be with me? (P16).* |
|  | It was difficult to work with patients with complex needs or have been convicted of a sexual offence (U) | *Then we expect them to behave differently, but without giving a single tool when it comes to sexuality and that part. They still have to understand the whole big thing, which is so difficult even for us who do not have an intellectual disability (P1).* |
|  | Sexuality being perceived as private and taboo makes silence easier than conversation (U) | *It’s just quiet about it, you, you just leave it out, until it becomes a problem. And then no one knows how to handle it and everyone, you easily fall into different morals. Opinion and much of one’s own morals and opinions and experiences are reflected in those discussions (P1).* |
|  | Patients were treated differently based on individual norms and values (E) | *No one did anything, it wasn’t addressed or discussed. Nor did he get an examination, he didn’t get any help, nothing, it was just swept under the carpet, yes, all the time, I experienced. I think it’s really disappointing (P14).* |
|  | Patients are sometimes seen by staff as having no need for a sexual life (U) | *We have a book trolley that goes around the wards, and as recently as yesterday we were told that a ward had removed a book called The Wankers. The staff who had to take the book back felt that this was just a title of the book, the book itself was not controversial in any way. We shouldn’t limit our patients’ access to literature, but still, the ward thought that this isn’t appropriate, so they had to take it back, it shouldn’t be on a library trolley in the ward, mmm, I feel THAT’S how we often work (P5).* |
|  | Staff felt that they lacked knowledge and competence concerning sexuality and sexual health (Un) | *N/A* |
|  | Staff were concerned about patients misinterpreting conversations and becoming sexually interested (U) | *Actually, I don’t know why we are so afraid to talk about it. Of course, it can be that you arouse or encourage things in the ward, and they shouldn’t have sexual relations with each other. If we have a permissive attitude maybe, and also have conversations about sexuality and sexual health, it might be interpreted that we have a permissive attitude. I don’t know if it that’s the case, or that it would increase the occurrence of relationships and vulnerability, I don’t know. I guess maybe that’s why we don’t talk about it (P5).* |
|  | Conversations about sexuality and sexual health need to be included in a structured way in forensic psychiatric care (U) | *The right tools or that we feel safe in the conversations and know how to talk about it. Yes, I just think it’s important and an important topic of conversation to include (P11).* |
|  | Staff expressed a need for increased knowledge and the development of an educational package containing general knowledge (U) | *How you can talk about, sometimes we have had, yeah, but supervision, where you can roleplay yourself, so in this like how, in different conversations, about how to have, and mainly in substance abuse there has been these difficult conversations, that you get to practice about how to have these conversations. Perhaps in the same way here, through supervision or that you get to practice, that you get to practice with each other, to have these types of conversations (P11).* |
|  | **Quinn, C., & Happell, B. (2012). Getting BETTER: Breaking the ice and warming to the inclusion of sexuality in mental health nursing care. *International Journal of Mental Health Nursing*, *21*(2), 154-162.** | |
|  | Discussing sexuality with service users is important to their identity and recovery (U) | *It’s really important (to talk about sexuality). It’s a really essential part of people’s identity, their recovery, and their wholeness as human beings, to have the ability to love, be loved, and to express that intimacy with someone else, and to value themselves. So I think it’s very important that we discuss this. (Shannelle)* |
|  | Training helped dispel fears of broaching a taboo subject (U) | *It was very useful. It just gave me the conﬁdence; it is important to ask. We do have the evidence to say we should ask; it is essential, and probably how to go about it.That was very important to me, because I didn’t know how to do it . . . with this model, we approach, we raise the topic, and we wait. We don’t need to have the answer. (Frank)* |
|  | Strong rapport and trust is conductive to conversations (U) | *Yeah, I think I’ll just include it. Not straight up; I’ll probably wait until the right time. I don’t think this is the typeof thing to talk about straight up, but perhaps a few days into the admission. I mean, if the guy is really psychotic,I wouldn’t bring it up, so I guess you need to pick the right time; once he’s well enough and the time is right. (Rhys)* |
|  | Training raised nurses' confidence in addressing sexuality (U) | *It’s like the stock standard now (including sexuality), andI’ve put it into my practice. I don’t get so red now myself talking about it. . . . I’ve found that by asking about possible sexual side-effects is a good way to get the conver-sation going. . . . it’s opened the door to the conversation.(Olivia)* |
|  | **Quinn, C., & Happell, B. (2015). Consumer sexual relationships in a Forensic mental health hospital: Perceptions of nurses and consumers. *International Journal of Mental Health Nursing*, *24*(2), 121-129.** | |
|  | Acknowledging service users as sexual beings (U) | *They are sexual beings and . . . they remain sexual beings,whether they are here for a week, 10 weeks, a year, or10 years. If anything, I would imagine it to become more depressing over time . . . and more frustrating watching the years go by and not having any outlet. . . . I just can’t imagine that. (Fiona)* |
|  | Intimate connections may reap therapeutic benefits (U) | *While a relationship in an institution might cause problems, it might also be very beneﬁcial . . . in the community they would be having the relationship, but it might be complicated by taking drugs. . . . So the fact here is that they have to go slow . . . so we are trying to support them in here and organize leaves together, so we are trying to watch the whole thing develop, support and nurture their relationship. (Alice)* |
|  | Staff's responsibility to monitor unhealthy relationships (U) | *For me, a concern is that we are working in a recovery-model setting where everything is focused on a progression through the rehab process, so how much do we see a relationship retarding a person’s progression? Relationships aren’t always a positive thing . . . this is a realistic concern. If it is a problem, when do we step in to helpout? (Greg)* |
|  | Danger of abuse or exploitation (U) | *I think. . . . there might be some danger of abuse of it or pressure in making a decision about being in a relationship or pressured to consent to sex or activity when they are not ready. I can think of some patients who might target particular vulnerable women or put the hard word on, to get involved in some type of sexual activity that they aren’t consenting to. (Ken)* |
|  | **Quinn, C., & Happell, B. (2015). Sex on show. Issues of privacy and dignity in a Forensic mental health hospital: Nurse and patient views. *Journal of clinical nursing*, *24*(15-16), 2268-2276.** | |
|  | Acknowledging service users' need for privacy and respect (U) | *Larry: One of my concerns is of sexual relationships occurring in public areas around the campus but then again there are no deﬁned areas for them to have some privacy so it’s a bit of a viscous circle that one. If they were having sex . . . in public areas there are cameras and the potential to be seen by other staff, patients or visitors.* |
|  | Risk management and safety as a key consideration (U) | *Alice: We did discuss as a team the need for this couple to have intimate time together and whether this should happen here in hospital or off campus in the community. There might be some advantage of this occurring out of the hospital. . . . so probably the location and no staff involvement are the two biggest things for me.* |
|  | Patients' sexual activity was only acceptable during "me time" (U) | *Danni: There is me time . . . we have curtains on the outside of the windows and have ‘modesty’ curtains on the inside so if they want some private time to masturbate they can put the curtain up and if you look through and see the curtain up well we would then knock and see if it is all right to open the door.* |
|  | **Quinn, C., & Happell, B. (2016). Supporting the sexual intimacy needs of patients in a longer stay inpatient forensic setting. *Perspectives in psychiatric care*, *52*(4), 239-247.** | |
|  | Concern for patients' capacity for consent (U) | *I think the assumption would be that for patients in acute,their mental state isn’t great and not stable and so they are in a state where they simply are unable to consent to be in a sexual relationship . . . they might be erratic or impulsive. . . their decision making processes are poor and all of these things contribute to problems in providing consent.* |
|  | High surveillance as a barrier to intimacy (U) | *I think it would be a little more difﬁcult in acute given that it is a much higher security area and where would you actually do it . . . there is obviously some type of high risk.* |
|  | The need for peer-developed guidelines (U) | *Whether a person is allowed or not to have sexual relationships they do occur no matter what. I guess by having a policy or some kind of formal way of identifying relationships would be a great step forward so things are occurring not in secrecy and we can support patients too, not by knowing exactly what they are doing, the details, but to support them and for them to know that they have our support.*  *I’d like to think that staff would be consulted. I think it is important to have staff involved, and their opinion matters, as opposed to some protocol developed and staffare asked to follow without consultation or discussion . . .* |
|  | Introduction of guidelines will be welcomed by patients (U) | *For the ones who are in relationships on campus it would be good because it would remove that element of sneaking around and doing it in places where human beings really shouldn’t be doing it and not having it be 'a dirty little secret'. They could actually do it, do it comfortably and do it properly and get their needs met.* |
|  | Clinicians' responsibility to support patient intimacy needs (U) | *I guess to a large extent that it may be the responsibility of the clinicians to ensure that people are supported. Not all friendships or relationships have their good days so it’s about how to educate and support patients around those issues during the relationship and how comfortable does the patient and their partner feel about having nurses sticking their noses into their relationships? . . . I guess this has a lot to do with the relationship and the level of rapport that the nurse has with the patient* |
|  | **Quinn, C., Happell, B., & Browne, G. (2011). Talking or avoiding? Mental health nurses' views about discussing sexual health with consumers. *International Journal of Mental Health Nursing*, *20*(1), 21-28.** | |
|  | Confidence in discussing sexuality varies with experience (U) | *think it’s just experience; I think I wouldn’t have been able to do that 25 years ago, maybe (Lisa).* |
|  | Issues surrounding sexuality should be explored after building rapport (U) | *Yes, I do (bring it up), but not always on the ﬁrst contact.Usually I have some degree of rapport with them (con-sumers) ﬁrst (Shannelle).* |
|  | Avoidance in discussing the topic (U) | *I think from a nursing point of view, it is still a taboo subject (Frank).* |
|  | The consumer is responsible for bringing up the topic first (E) | *If someone brings it up as an issue that is really affecting them, then you know, I’d make time to sit down and listen to what they have to say (Louise).* |
|  | Sexual concerns of patients are a lesser priority (U) | *I guess it goes back to . . . (the) other issue(s) at hand that need addressing, and that (sexual assessment) gets pushed down to the bottom. . . . It’s not as important as their experience (of) side-effects and symptoms of their mental illness. I guess I consider that I have greater expertise in helping them with symptoms of their mental illness, rather than talking about and assisting them with their sexual issues (Louise)* |
|  | Nurses were not encouraged to discuss sexuality in assessments (U) | *The funny thing is when doing assessments, nowhere on the assessment sheet does it say anything about sexuality.There are no questions; there is nothing (asking) ‘Are you sexually active?’ Nothing. It avoids it too (laughs); we avoid it like the plague (laughs) (Jean).* |
|  | Sexual concerns are not the nurses' areas of expertise (U) | *I don’t see myself as an expert in that department. It’s often the case that I’ll say we’ll talk about it later, at your next doctor’s appointment . . . he’s the one who often answers these kind of questions (Louise).* |
|  | Nurses' discomfort with patients' right to sexuality (U) | *When you talk about sexuality, a large number of staff tend to be very uncomfortable with the idea; they believe that our patients should be asexual, they don’t have a right to sexuality, and they feel very uncomfortable that these people should have a sexual life. . . . It is as if people with a mental illness have no right to sexuality (Ethan).* |
|  | **Raisi, F., Yahyavi, S., Mirsepassi, Z., Firoozikhojastefar, R., & Shahvari, Z. (2017). Neglected sexual needs: A qualitative study in Iranian patients with severe mental illness. *Perspectives in psychiatric care*, *54*(4), 488-494.** | |
|  | Lower priority of sexual issues for clinician at the first interview (U) | *“Duration of admission in acute wards is usually 4–6 weeks and in this period of time, symptoms remission is very important, so sexual health problems are in the second rank”(Participants code 4).* |
|  | Clinicians‘ attitude and cultural barriers (U) | *“In our community, talking about sexual issues is very difficult for clinicians. How could a clinician educate patients regarding sexual issues when he/she does not feel comfortable in talking about these concerns? In psychotherapy, the patient does not speak about a subject that is difficult for the therapist. Clinicians should ask some questions about sexual concerns for helping their patients; however, they are uncomfortable with these issues” (Participants code 6, a 59-year-old clinician).* |
|  | Poor understanding of sexual problems management (U) | *“The patients’ sexual needs are completely ignored by the healthcare providers; because we believe that they do not have sexual needs. Patients are prone to risk taking sexual behaviors, despite decreased libido due to medications side effects.”(Participants code 2)* |
|  | **Southall, D. J., & Combes, H. A. (2022). Clinical psychologists’ views about talking to people with psychosis about sexuality and intimacy: A Q-methodological study. *Sexual and Relationship Therapy*, *37*(4), 512-536.** | |
|  | Sexuality as a fundamental human issue (U) | Participant 17 highlighted the ‘*need to acknowledge the relevance and importance of sex and intimacy in people’s lives.We can’t pretend it doesn’t exist*’. |
|  | Conversations around intimacy can help achieve goals of therapy (U) | Participant 15 highlighted that *‘In order for therapy to be effective we ought not to occlude any areas of normal human life, including sex and intimacy’*. |
|  | Patients may find it challenging to discuss intimacy (U) | Participant 16 supported this with the statement ‘*sex is not something we generally talk about publicly as we might some other things, so we have to recognise that this might be particularly embarrassing, more or less so depending on their own reference points’*. |
|  | Discussions may cause otherwise avoidable negative consequences (U) | Participant 14 commented: “*If my client misinterpreted my interest this would disrupt the therapeutic alliance considerably and may contribute to complex transference*.” |
|  | Whether clinical psychologists have sufficient training about raising discussions around intimacy (U) | Doctoral training should provide clinical psychologists ‘*with the ability to talk about every subject relevant to human experience. Sex is just another aspect of human experience*’ (Participant 11).  ‘for such a complex and immense topic, there was very little in the way of training…’. (Participant 13) |
|  | Considerations of risk to the patient (U) | Participant 4 stated ‘*I’d need to if it was part of the client’s difficulties or increased their risk profile in some way*’. |
|  | Intimacy is only addressed if the client wishes to discuss (U) | Participant 13 commented *‘It is generally a topic which seems to be avoided by other disciplines in mental health services and yet it is a significant part of people’s lives. Someone has to be available to talk with clients about these things if they wish to do so’.* |
|  | Sex and intimacy are not part of their role (U) | *‘it is not necessarily part of my role’ (Participant 12).* |
|  | **Tennille, J., Bohrman, C., Barrenger, S., Compton, E., Meduna, E., & Klein, L. (2022). Behavioral health provider attitudes and beliefs about sexuality and intimacy: findings from a mixed method design. *Community Mental Health Journal*, *58*(3), 444-453.** | |
|  | Increasing confidence around conversations (Un) | Not Supported. |
|  | Need for information around sexuality and intimacy (U) | “*need to understand the physical, physiological, social and emotional factors associated with and reflecting sexuality and intimacy*” |
|  | Training increases comfort and decreases stigma (U) | *“Learning this information among other committed professionals who are also inexperienced, taught by truly relaxed and non-judgmental experienced clinicians really helps dissolve discomfort!”* |
|  | Need for support from the agency (U) | *"When working for an agency, the support of the leadership is crucial.”* |
|  | Need for information around working with LGBT individuals (U) | *Another mentioned wanting “increased knowledge related to how various populations engage in sexual activity, outside of cisgender, heterosexual individuals.”* |
|  | **Tiwana, R., McDonald, S., & Völlm, B. (2016). Policies on sexual expression in forensic psychiatric settings in different European countries. *International journal of mental health systems*, *10*, 1-11.** | |
|  | Sexual expression as a human right (U) | *“Sexual expression is an important part of human nature” (Netherlands)* |
|  | Viewing sexual expression in relation to risk management (U) | *“Too much repression…can even lead to exaggerated frustration and elevation of risk levels” (Netherlands).* |
|  | Focus on the biological aspect of sexual expression (U) | *“…should we issue condoms to male patients if we feel they’ve got a relationship with somebody with the same gender? But we decided that that just opened too much of a floodgate really…” (UK, high secure)* |
|  | The hospital as an inappropriate setting for intimate relations (U) | *“…it’s a public space so that’s why anything of intimacy would be discouraged and not acceptable” (UK, high secure)* |
|  | Public opinion that patients' freedom for sexual expression should be curtailed (U) | *“…public opinion is that they have a very good life in prisons and in hospitals which is not true…so now you even allow them to have sex?” (Switzerland)* |
|  | Rigid guidelines are unfavoured by patients (U) | *“… his girlfriend came to visit and they were a bit too intimate in front of the staff … and they were discouraged from this intimacy and she was dressed inappropriately as well and we put a stop to that, and the patient challenged that and said it’s my girlfriend” (UK, high secure)* |
|  | Staff find it difficult to discuss sex (U) | *“Staff sometimes find it hard because they find it hard to discuss sex as a topic, particularly homosexual relations, but this reluctance can very well be explained by their personal feelings concerning homosexuality” (Netherlands)* |
|  | A need for policy change (U) | “I was unaware of the extent of the conjugal visits in Europe and it might be something we should debate more openly” (UK, medium secure) |
|  | **Tumwakire, E., Arnd, H., & Gavamukulya, Y. (2022). A qualitative exploration of Ugandan mental health care workers’ perspectives and experiences on sexual and reproductive health of people living with mental illness in Uganda. *BMC Public Health*, *22*(1), 1722.** | |
|  | People with mental illness have normal sexuality needs (U) | *“We should know that being mentally sick doesn’t take away your sexual feelings, it doesn’t. these are normal women, these are normal men and if they see this young musawo looking nice they will definitely give you sexual advances.” (p7)* |
|  | Desire to maintain relationships as motivation for treatment adherence (U) | *“You normally find that such people they comply to their treatment very well for security of their marriage.” (p10)* |
|  | Staff experiences of sexual assault are a barrier to care (U) | *“We don’t want to work alone especially over the weekend, because some patients have high libido, they can rape you.” (p2)* |
|  | Family planning services as a solution (U) | *“Yes, they come here and they involve themselves in sex so eventually it became too much in the compounds and in the meeting we had to decide to administer family planning to them without their consent without their choice. It is now us the health workers basing on the observation we have made to decide what to do for them but not giving them that opportunity.” (p12)* |
|  | Sexual and reproductive health is outside of the mental health specialty (U) | *“For us here in the mental clinic we may not dwell so much on the sexual problem, we usually dwell mostly on the mental problems, hallucinations etc.” (p11)* |
|  | Limited knowledge and training about sexuality needs (U) | *“We usually have CMES every week at least but those CMEs are usually on mental health and never on reproductive health.” (p7)* |
|  | Heavy workload prevents staff from addressing intimacy (U) | *“The clinic is usually heavy with very many patients. Unless if the patient raises it specifically that is when you can go into that.” (p8).* |
| **Urry, K., Breakey, G. R., Scholz, B., & Chur‐Hansen, A. (2023). Approaches for improving sexuality and sexual health care in mental health settings: A qualitative study exploring clinicians' own perspectives. *International Journal of Mental Health Nursing*, *33*(1), 125-133.** | | |
| The need for more sexuality-related education and training (U) | | *I think for people in the health system, [and] certainly mental health that are time poor and having requirements to do everything else [… we should have] an online, course about sexual health and what it means and how to approach it with clients and ideas and tools to use and stuff, that it can be done in half an hour an hour session of online training would be really helpful, online. I think it would be far less intrusive or confronting for people to have to do that online than to sit in a room with people. (Mary, nurse)* |
| The need for clear information on referral pathways (U) | | *I don't feel like I have a good handle on who would be a good, safe referral pathway […] you know, if I sent someone who was struggling with their sexuality [identity] or struggling with reckless sexual contact and wanted a really kind of non-judgmental approach to that, I wouldn't have a good sense [of where to refer them]. (Melissa, psychologist)* |
| The need for support from both direct peers and colleagues (U) | | *Interviewer: Yeah. Is it something you would talk about with colleagues or in peer supervision or anything like that?  Stephanie: Yeah, sometimes, but not as much as I would like to. I do a lot of—in my private practice—do a lot of couple stuff, so, sex comes up a fair bit, um. But I haven't got—had a lot of supervision (Stephanie, psychologist)* |
| There were specific structural challenges or barriers, time constraints that made it challenging to prioritize sexuality-related needs (U) | | *I think I did send an email, out into the ether, after we talked about it [a particular issue at the previous interview…] like, "Oh, there's a thing that has to be changed," and I don't think I've ever heard anything back about it. I might have even, I guess raised [at] a… too small of a meeting for it to have an impact. I think I might've talked to my team later about it and she said, "That sounds like a good idea," and it never went anywhere. (Melissa, psychologist)* |
| The importance of clinicians building comfort with thinking or talking about, and addressing, sexuality-related needs within their clinical practice (U) | | *I was teaching somebody to do quality of life, and it talks about orgasms and things like that, and this [junior nurse] (chuckle) you see her hesitate and I said, “Look, you gotta get over that” […] you can't look embarrassed by asking a question or hesitate while you're doing it. You got to prove to that person that, whatever they say is okay. (Tina, nurse)* |
| Staff should actively reflect on their own experiences, beliefs, values to facilitate improved sexuality-related practice (U) | | *“I think it is important to reflect on your own judgements, because […] we've all got our own experiences, or thoughts around this and it does influence what you do.” (Stephanie, psychologist).* |
|  | **Urry, K., Chur‐Hansen, A., & Khaw, C. (2019). ‘It's just a peripheral issue’: A qualitative analysis of mental health clinicians’ accounts of (not) addressing sexuality in their work. *International Journal of Mental Health Nursing*, *28*(6), 1278-1287.** | |
|  | Sexuality is hard to talk about (U) | *and people being embarrassed and not willing to or not necessarily identifying that so you need to ask [about treatment-induced sexual dysfunction. . .] I think people wouldn’t necessarily bring it up. Or they’d be reluctant until they know you better or whatever. (Eric,psychiatrist)* |
|  | It is service users' responsibility to raise conversations around sexuality (U) | *I don’t really see [sexual concerns] as something that Iwould necessarily raise because I’m not setting the agenda. They are. They come to me with particular presenting complaints and I’m responsive to that [. . .](Josh, psychiatrist)* |
|  | Clinicians' comfort with their own sexuality and intimacy needs (U) | *I think if people aren’t okay with their own sexuality their own. . . comfortableness with themselves their own self-esteem all of that stuff has an impact on whether you’re able to talk to clients about [sexuality and sexual health. . .] (Yvonne, nurse)* |
|  | Sexuality does not belong in a mental health setting (U) | *We’re probably not skilled or trained or educated enough to pass that [kind of information] onto the young people. It’s kind of just like a peripheral issue for us [. . .] We note [sexuality-related concerns] but we don’t directly deal with it. (Dean, nurse)* |
|  | Clinicians' focus on sexual health and risk rather than intimacy and relationships (U) | *Well obviously our mental health patients are the most vulnerable in terms of sexual health because a lot of our clients will either have sex for money or drugs and not have protected sex and they’ll have sex with a lot of different men. A lot of our consumers are so vulnerable they get raped and you know obviously are too scared to tell anyone because it re-traumatises them. (Jake,nurse)* |
|  | Intimacy needs are a peripheral issue (U) | *when someone comes in here often they’re trying to hurt themselves or others or putting themselves or others at risk in some way and that tends to trump you know ‘how’s your relationship going how your sex life going are you having any problems with that’, that seems to come later um and in a public system [. . .]you may only be seeing people in those times of crisis so you might not be building the relationship up to remember to ask to think to ask to have time to ask about all of those other factors [. . .] (Emma, psychia-trist)* |
|  | Sexuality should be part of the recovery process (U) | *I think it’s probably something that we don’t ask about that much. Probably as a team. Or I’ve worked with a couple of different teams so it’s not just this team butas a public mental health system [. . .] I think we don’t talk about sexuality as much as we should when we’re assessing and treating. (Fay, psychologist)* |
|  | **White, R., Haddock, G., & Varese, F. (2019). Supporting the intimate relationship needs of service users with psychosis: What are the barriers and facilitators?. *Journal of Mental Health*.** | |
|  | Stigma against the capacity for romantic relationships in people with psychosis (U) | *People who I work with who are in relationships…those relationships are quite difficult for them and do cause them a lot of distress, sort of either on-off relationships or partners not being able to cope with the symptoms that service users are experiencing. (P17)* |
|  | Risk management as a priority (U) | *I think we’re probably more concerned about vulnerability issues – whether people are being exploited in any way – so they may get with a partner who may exploit them for money or other things. (P15)* |
|  | Lack of guidance on the boundaries of their role (U) | As a result participants perceived that the boundaries of their role as a professional were unclear: “*Appropriate boundaries and how far to take that kind of support and treatment, because I think that’s something that’s very difficult to decipher”* (P4). |
|  | Intimacy needs related to recovery process (U) | *Having romantic partners, what comes with that is a lot of protective factors; the understanding, the support, the sounding out their worries and normalisation. (P20)* |
|  | A good therapeutic relationship is conductive to discussions (U) | *I don’t know, maybe it is just because I’m [a] similar age and they feel that maybe [it’s] less embarrassing to talk about with me than it is with somebody who’s like their mum. (P17)* |
|  | Desire for guidance and training (U) | “*I think it could be guidance policy. What’s appropriate? What’s our clinical role in that? The expectation*.” (P18). |
|  | Onus on staff with social roles to address intimacy needs (U) | *“You stick to what you’re good at and you can maybe give ‘em a bit of advice but really you’re better off signposting to somebody who deals in that all day” (P19).* |

**Appendix D**

***ConQual Assessment protocol***

**The ConQual approach**

The ConQual approach presents a grading of confidence in each synthesised finding. Each synthesised finding begins with a ranking of high confidence and this is downgraded or remains the same depending on the dependability and credibility of the findings that make up that finding. If the majority of findings in a synthesised finding come from articles rated ‘high’ in their dependability then the ranking of high confidence remains the same. If the majority of findings within a category come from articles that are ‘moderate’ or ‘low’ in their dependability then the category is downgraded from ‘high’ confidence to ‘moderate’ or ‘low’. The confidence ranking is then altered again depending on the credibility of the findings within the synthesised finding. If the category is composed of findings supported by only ‘Unequivocal’ illustrations, then the ranking remains the same. If the category is composed of a mixture of ‘Unequivocal’ and ‘Equivocal’ findings the category is downgraded by one level. If the category is composed of totally ‘Equivocal’ findings or a mixture of ‘Unequivocal’, ‘Equivocal’ and ‘Unsupported’ findings its ranking is downgraded two levels.

**Dependability**

When rating the dependability of an article the reviewer asks five questions:

1. Is there congruity between the research methodology and the research question or objectives? 2. Is there congruity between the research methodology and the methods used to collect data?

3. Is there congruity between the research methodology and the representation and analysis of data?

4. Is there a statement locating the researcher culturally or theoretically?

5. Is the influence of the researcher on the research, and vice-versa, addressed?

By answering yes or no to each question each study is given a score out of five. One reviewer (EW) assessed the dependability of each study independently using the JBI Checklist for Qualitative Research. Studies scoring 4 or 5 are given a ranking of high dependability. Studies scoring two or three are given a ranking of moderate dependability. Studies scoring one or zero are given a ranking of low dependability.

**Credibility**

When assessing credibility the focus is not on the research study itself but on the individual findings within an article and whether there is sufficient evidence to support each claim the author makes. On data extraction each finding is accompanied by an ‘illustration’ or a piece of evidence that may be a quotation from a participant or field notes. The credibility of each finding is rated as ‘unequivocal’ where the finding is not open to challenge due to the accompanying illustration, ‘equivocal’ where the accompanying illustration lacks clear association with the finding and is therefore open to challenge, or ‘unsupported’ where there is no clear accompanying illustration.

**Appendix E**

***Excluded studies***

The following studies did not meet the inclusion criteria during full-text screening and were excluded from the final review. A hierarchy of exclusion reasons was consulted for studies that presented with multiple reasons for exclusion.

Hierarchy of exclusion reasons:

1. Wrong study design
2. Wrong setting
3. Wrong patient population

Abbott, D., & Howarth, J. (2007). Still off‐limits? Staff views on supporting gay, lesbian and bisexual people with intellectual disabilities to develop sexual and intimate relationships?. *Journal of Applied Research in Intellectual Disabilities*, *20*(2), 116-126.

**Reason for exclusion:** wrong patient population

Achey, N. (2020). *Direct support professionals' perspectives on sexuality issues of adults with intellectual disabilities: A qualitative analysis of interviews with providers in Maine*. The University of Maine.

**Reason for exclusion:** wrong patient population

Alston, L. (2018). *Licensed Therapists' Work with Sexual Minority Victims of Intimate Partner Violence: A Generic Qualitative Study* (Doctoral dissertation, Capella University).

**Reason for exclusion:** wrong patient population

Bates, C., McCarthy, M., Milne Skillman, K., Elson, N., Forrester‐Jones, R., & Hunt, S. (2020). “Always trying to walk a bit of a tightrope”: The role of social care staff in supporting adults with intellectual and developmental disabilities to develop and maintain loving relationships. *British Journal of Learning Disabilities*, *48*(4), 261-268.

**Reason for exclusion:** wrong patient population

Carnaby, S., & Cambridge, P. (2002). Getting personal: an exploratory study of intimate and personal care provision for people with profound and multiple intellectual disabilities. *Journal of Intellectual Disability Research*, *46*(2), 120-132.

**Reason for exclusion:** wrong patient population

Charitou, M., Quayle, E., & Sutherland, A. (2021). Supporting adults with intellectual disabilities with relationships and sex: A systematic review and thematic synthesis of qualitative research with staff. *Sexuality and Disability*, *39*, 113-146.

**Reason for exclusion:** wrong patient population

Chrastina, J., & Večeřová, H. (2020). Supporting sexuality in adults with intellectual disability—a short review. *Sexuality and Disability*, *38*(2), 285-298.

**Reason for exclusion:** wrong setting

Ćwirynkało, K., Byra, S., & Żyta, A. (2017). Sexuality of adults with intellectual disabilities as described by support staff workers. *Hrvatska revija za rehabilitacijska istrazivanja*, *53*, 77-87.

**Reason for exclusion:** wrong setting

Deffew, A., Coughlan, B., Burke, T., & Rogers, E. (2022). Staff member's views and attitudes to supporting people with an intellectual disability: a multi‐method investigation of intimate relationships and sexuality. *Journal of Applied Research in Intellectual Disabilities*, *35*(4), 1049-1058.

**Reason for exclusion:** wrong setting

Ettalibi, M. Y., Marchi, M., Magarini, F. M., Landi, G., Mattei, G., Pingani, L., & Galeazzi, G. M. (2019). Affective and sexual needs of residents in psychiatric facilities. *Minerva Psichiatrica*.

**Reason for exclusion:** wrong study design

Evans, D. S., McGuire, B. E., Healy, E., & Carley, S. N. (2009). Sexuality and personal relationships for people with an intellectual disability. Part II: staff and family carer perspectives. *Journal of Intellectual Disability Research*, *53*(11), 913-921.

**Reason for exclusion:** wrong patient population

Fonseca, M. I., Almeida, D., Martins, A. P., Cerqueira, M., Villar, F., Martinez de Oliveira, J. M., & Afonso, R. M. (2022). Sexual expression involving people with dementia living in long‐term care facilities: Staff's reactions. *International Journal of Older People Nursing*, *17*(6), e12474.

**Reason for exclusion:** wrong patient population

Ikebuchi, E. (2015). Supporting the love, marriage, and child-rearing of persons with schizophrenia. *Seishin Shinkeigaku Zasshi= Psychiatria et Neurologia Japonica*, *117*(11), 910-917.

**Reason for exclusion:** wrong study design

Grieve, A., McLaren, S., Lindsay, W., & Culling, E. (2009). Staff attitudes towards the sexuality of people with learning disabilities: a comparison of different professional groups and residential facilities. *British Journal of Learning Disabilities*, *37*(1), 76-84.

**Reason for exclusion:** wrong setting

Holler, R., & Bondorevsky-Heyman, C. (2024). The Dynamics of Intimate Relations in Residential Settings for People with Intellectual Disabilities: Social Workers’ Perspective. *Sexuality Research and Social Policy*, *21*(1), 422-435.

**Reason for exclusion:** wrong setting

Ishay, G. (2018). Relationships and Sexuality in Occupational Therapy - Occupational Therapy Group Intervention for Young Adults with Psychiatric Disorders: A Case Study. Israeli Society of Occupational Therapy, 27(2), E55

**Reason for exclusion:** wrong setting

Lam, A., Yau, M. K., Franklin, R. C., & Leggat, P. A. (2022). Challenges in the delivery of sex education for people with intellectual disabilities: A Chinese cultural‐contextual analysis. *Journal of Applied Research in Intellectual Disabilities*, *35*(6), 1370-1379.

**Reason for exclusion:** wrong patient population

Lichtenberg, P. A. (2014). Sexuality and physical intimacy in long-term care. *Occupational therapy in health care*, *28*(1), 42-50.

**Reason for exclusion:** wrong setting

Lines, J., Combes, H., & Richards, R. (2021). Exploring how support workers understand their role in supporting adults with intellectual disabilities to access the Internet for intimate relationships. *Journal of Applied Research in Intellectual Disabilities*, *34*(2), 556-566.

**Reason for exclusion:** wrong patient population

Luby, R. (2019). How one nurse helped break the taboo of talking about sex in mental healthcare settings. *Mental Health Practice*, *22*(4).

**Reason for exclusion:** wrong study design

Morales, G. E., Lopez, E. O., & Mullet, E. (2011). Acceptability of sexual relationships among people with learning disabilities: family and professional caregivers’ views in Mexico. *Sexuality and Disability*, *29*, 165-174.

**Reason for exclusion:** wrong setting

Nørtoft, K., & Rubin, S. E. (2024). Let’s Talk About Sex! Perspectives from People with Intellectual and Developmental Disabilities and Caregivers in Residential Institutions in Greenland. *Sexuality and Disability*, *42*(2), 225-242.

**Reason for exclusion:** wrong patient population

Oloidi, E. O., Northway, R., & Prince, J. (2022). ‘People with intellectual disabilities living in the communities is bad enough let alone… having sex’: Exploring societal influence on social care workers' attitudes, beliefs and behaviours towards support for personal and sexual relationship needs. *Journal of applied research in intellectual disabilities*, *35*(4), 1037-1048.

**Reason for exclusion:** wrong setting

Piantedosi, D. K., Reed, K., & O'Shea, A. (2023). Supporting occupational therapists to initiate conversations about sexuality with people with intellectual disability: Co‐design by deliberative dialogue. *Australian Occupational Therapy Journal*, *70*(5), 581-598.

**Reason for exclusion:** wrong setting

Prountzos, T. (2023). *The Experiences of Women With Intellectual Disabilities Around Sex and Intimacy: A Thematic Synthesis Clinical Psychologists’ Experiences of Exploring Sex and Intimacy With People With Intellectual Disabilities and Their Networks: A Phenomenological Study* (Doctoral dissertation, Canterbury Christ Church University (United Kingdom)).

**Reason for exclusion:** wrong setting

Schouten, V., Henrickson, M., Cook, C. M., MacDonald, S., & Atefi, N. (2023). Value pluralism about sexual intimacy in residential care. *Nursing ethics*, *30*(3), 437-448.

**Reason for exclusion:** wrong setting

Sung, S. C., Lin, Y. C., Hong, C. M., & Cho, P. P. (2007). An exploratory study of psychiatric patients' needs and nurses' current practices related to sexual counseling. *Hu li za zhi The Journal of Nursing*, *54*(1), 43-52.

**Reason for exclusion:** wrong study design

Van Son-Schoones, N., & Van Bilsen, P. (1995). Sexuality and autism: a pilot-study of parents, health-care workers and autistic persons. *International Journal of Adolescent Medicine and Health*, *8*, 87-102.

**Reason for exclusion:** wrong setting

Wright, D. E. (2006). Predictors of sex-related discussions between treatment staff and clients with severe mental illness.

**Reason for exclusion:** wrong study design

Wright, D., & Pugnaire-Gros, C. (2010). Let's talk about sex: Promoting staff dialogue on a mental health nursing unit. *Journal for Nurses in Professional Development*, *26*(6), 250-255.

**Reason for exclusion:** wrong study design

Young, R., Gore, N., & McCarthy, M. (2012). Staff attitudes towards sexuality in relation to gender of people with intellectual disability: A qualitative study. *Journal of Intellectual and Developmental Disability*, *37*(4), 343-347.

**Reason for exclusion:** wrong setting

**Appendix F**

***Findings contributing to each category***

Please refer to Appendix C for the originating study of each finding.

| Category | Finding |
| --- | --- |
| Universality of intimacy needs | Service users' sexual behaviours are no different than ours (U) |
|  | Acknowledging service users as sexual beings (U) |
|  | Sexuality as a fundamental human issue (U) |
|  | Sexual expression as a human right (U) |
|  | People with mental illness have normal sexuality needs (U) |
|  | It is important to normalise sexuality (U) |
|  | Conversations around sexuality felt natural and like any other subject (U) |
|  | Topics surrounding sex and sexuality were engaging and important to work with (U) |
| Distinction between sexual and intimacy needs | Education programs focus on sexual health instead of intimate relationships (U) |
|  | Focus on the biological aspect of sexual expression (U) |
|  | Clinicians' focus on sexual health and risk rather than intimacy and relationships (U) |
|  | Desire for romantic relationships is seen as a legitimate recovery-related goal, while purely sexual needs are not (U) |
| Stigma against people with mental illness | Stigma against intimacy needs in patients with mental illness (U) |
|  | Societal stigma against people with mental illness dating (U) |
|  | Public opinion that patients' freedom for sexual expression should be curtailed (U) |
|  | Families being overprotective (U) |
|  | Patients are sometimes seen by staff as having no need for a sexual life (U) |
| Considerations of the service users' capacity for relationship building and maintenance | Family planning is encouraged (U) |
|  | Family planning services as a solution (U) |
|  | Stigma against service users' ability to form intimate relationships (U) |
|  | Belief that relationships are inappropriate for the service user (U) |
|  | Concern for patients' capacity for consent (U) |
|  | Stigma against the capacity for romantic relationships in people with psychosis (U) |
|  | Doubts about service users' ability to communicate their needs (U) |
|  | Patients may find it difficult to disclose mental illness to intimate partners (U) |
|  | Patients may find it challenging to discuss intimacy (U) |
|  | It was difficult to work with patients with complex needs or have been convicted of a sexual offence (U) |
|  | Parenthood was viewed as a controversial topic (U) |
| Intimacy needs are not prioritised in mental health settings | Sexual concerns of patients are a lesser priority (U) |
|  | Lower priority of sexual issues for clinician at the first interview (U) |
|  | Intimacy needs are a peripheral issue (U) |
|  | Addressing sexual health was less of a priority in clinical practice (U) |
| Confidence of staff | Lack of training to discuss sexuality and intimacy issues (U) |
|  | Lack of knowledge in how to deliver support (U) |
|  | Limited knowledge and training about sexuality needs (U) |
|  | Lack of guidance on the boundaries of their role (U) |
|  | Lack of training on how to discuss intimacy (U) |
|  | Poor understanding of sexual problems management (U) |
|  | Lack of confidence in addressing needs for intimacy (U) |
|  | Feelings of discomfort, loneliness or resourcelessness related to a perceived inability to provide adequate support (U) |
|  | Confidence in discussing sexuality varies with experience (U) |
|  | Staff felt a lack of knowledge and confidence in broaching the subject (U) |
|  | It was difficult to differentiate between safeguarding issues and acceptable lifestyle choices (U) |
|  | Staff were aware of sexual health needs of service users (U) |
|  | Staff expressed powerlessness as they didn't know how the patient can be best-helped (E) |
| Staff's discomfort and avoidance with the topic of intimacy | Discomfort with discussing sex (U) |
|  | Avoidance of and discomfort with discussing sex with service users (U) |
|  | Avoidance in discussing the topic (U) |
|  | Staff find it difficult to discuss sex (U) |
|  | Sexuality is hard to talk about (U) |
|  | Nurses' discomfort with patients' right to sexuality (U) |
|  | Relationship seeking support is perceived as inappropriate (U) |
|  | Sexuality being perceived as private and taboo makes silence easier than conversation (U) |
|  | The importance of clinicians building comfort with thinking or talking about, and addressing, sexuality-related needs within their clinical practice (U) |
|  | There was a fear of asking questions which were too private or uncomfortable (U) |
|  | The topic of sexuality was perceived to be difficult and challenging (U) |
|  | Staff were sometimes caught off guard when engaging in conversations around sex and sexuality (U) |
| Self-reflections on experiences and beliefs relating to intimacy | Clinicians‘ attitude and cultural barriers (U) |
|  | Clinicians' comfort with their own sexuality and intimacy needs (U) |
|  | Staff engaged in self-reflection about different situations they encountered (U) |
|  | Patients were treated differently based on individual norms and values (E) |
|  | Staff should actively reflect on their own experiences, beliefs, values to facilitate improved sexuality-related practice (U) |
|  | Staff reported a range of emotional experiences based on their experience of control over the conversation (U) |
|  | Clinicians‘ attitude and cultural barriers (U) |
| Having the right tools | Need for improved knowledge, training, and access to an integrated service provision (U) |
|  | Need for information around sexuality and intimacy (U) |
|  | Need for information around working with LGBT individuals (U) |
|  | Clinical psychologists have sufficient training about raising discussions around intimacy (U) |
|  | Training helped dispel fears of broaching a taboo subject (U) |
|  | Training raised nurses' confidence in addressing sexuality (U) |
|  | Training increases comfort and decreases stigma (U) |
|  | Staff expressed a need for increased knowledge and the development of an educational package containing general knowledge (U) |
|  | The need for more sexuality-related education and training (U) |
|  | The need for clear information on referral pathways (U) |
|  | There was a need for training for all staff groups (U) |
| The displacement of responsibility to address intimacy needs | Uncertainty about whether intimacy falls within their scope of work (U) |
|  | Sexual concerns are not the nurses' areas of expertise (U) |
|  | Sexual and reproductive health is outside of the mental health specialty (U) |
|  | Sexuality does not belong in a mental health setting (U) |
|  | Onus on staff with social roles to address intimacy needs (U) |
|  | Finding a relationship is beyond staff's scope of work (U) |
|  | Sex and intimacy are not part of their role (U) |
|  | Intimacy is only addressed if the client wishes to discuss (U) |
|  | The consumer is responsible for bringing up the topic first (E) |
|  | It is service users' responsibility to raise conversations around sexuality (U) |
|  | Sexual health provision in mental health services was limited (E) |
|  | Patients who were open to broaching difficult subjects were seen as facilitating the work of the staff (U) |
|  | Conversations should take place on patients' terms and their own language (U) |
| The importance of nurturing and respecting the therapeutic relationship | A good therapeutic relationship is conductive to discussions (U) |
|  | Issues surrounding sexuality should be explored after building rapport (U) |
|  | Strong rapport and trust is conductive to conversations (U) |
|  | Staff's responsibility to monitor unhealthy relationships (U) |
|  | Power differentials between provider and patient were a barrier (E) |
|  | It would be unethical to help service users find a relationship (U) |
|  | Staff were careful to maintain a professional distance from patients (U) |
|  | Staff felt a need to set boundaries in conversations with patients (U) |
|  | Concerns arose when the professional therapeutic relationship shifted to something more private (U) |
| Addressing intimacy needs as part of recovery-oriented practice | Importance of recovery-oriented practice (U) |
|  | Having conversations encourages recovery (U) |
|  | Discussing sexuality with service users is important to their identity and recovery (U) |
|  | Sexuality should be part of the recovery process (U) |
|  | Intimacy needs related to recovery process (U) |
|  | Support regarding intimate relationships is relevant to service users (U) |
|  | Intimate connections may reap therapeutic benefits (U) |
|  | Desire to maintain relationships as motivation for treatment adherence (U) |
|  | Clinicians' responsibility to support patient intimacy needs (U) |
|  | Conversations around intimacy can help achieve goals of therapy (U) |
|  | Patients often benefit from having sexuality explicitly discussed (E) |
|  | It was important for mental health professionals to address sexual health as part of their provision of holistic care (U) |
|  | Therapeutic processes should respond to the individual need of patients (U) |
|  | The experience of conversations often revolved around trying to understand the patient (U) |
|  | Staff adopted a listening approach and allowed patients to take control of conversations (U) |
| Concerns about the impact on service users' mental health | Service users must first be stable (U) |
|  | Viewing service users via a psychiatric lens (U) |
|  | Discussions may cause otherwise avoidable negative consequences (U) |
|  | Conversations may cause patients to deteriorate (U) |
|  | Staff were concerned about causing distress to the service user and damaging the therapeutic alliance (U) |
|  | Prostitution was predominantly viewed with skepticism (U) |
| Restrictive clinical setting | Sexual activity was strictly policed by clinicians and in accordance with the ‘policy’ (U) |
|  | Service users' sexual activity is highly scrutinised (U) |
|  | High surveillance as a barrier to intimacy (U) |
|  | Policy as a way to secure harmonious therapeutic environment (E) |
|  | Patients' sexual activity was only acceptable during "me time" (U) |
|  | The hospital as an inappropriate setting for intimate relations (U) |
|  | Patients experience a lack of privacy, sometimes due to overcrowding (U) |
|  | Visitation areas do not explicitly enable or promote intimacy (U) |
|  | Satisfaction of sexual needs was perceived to be harder for patients with longer lengths of stay (U) |
| Prioritising risk management and safeguarding | Romantic relationship needs are only addressed if there are safeguarding concerns (E) |
|  | Risk management and safety as a key consideration (U) |
|  | Viewing sexual expression in relation to risk management (U) |
|  | Risk management as a priority (U) |
|  | Danger of abuse or exploitation (U) |
|  | Considerations of risk to the patient (U) |
|  | Sexual decisions that affect others are seen differently (U) |
|  | Providers are exposed to awkward situations relating to patient's sexuality (U) |
|  | Staff experiences of sexual assault are a barrier to care (U) |
|  | Conversations around sexuality are seen as risk reducing in terms of both compliance to treatment and potential relapse (U) |
|  | The mission to protect the society is often prioritized higher than patients’ individual needs and rights for care (U) |
|  | Staff were concerned about patients misinterpreting conversations and becoming sexually interested (U) |
|  | Staff felt a professional responsibility to guide and protect the patient (E) |
|  | Sexuality was only taken into account if there were concerns such as the risk of pregnancy or abuse (U) |
|  | Service users who were perceived as being more high risk were more likely to receive sexual health care (U) |
|  | A careful approach to sexuality is needed for sex offenders (U) |
| Counterproductive policies | Program-level policies are counterproductive to navigating client goals to seuxality (U) |
|  | Restricting intimate relations on-site endangers the safety of patients (U) |
|  | Nurses were not encouraged to discuss sexuality in assessments (U) |
|  | A need for policy change (U) |
|  | Vague and arbitrary guidelines on 'inappropriate sexual behaviours' (E) |
|  | There were specific structural challenges or barriers, time constraints that made it challenging to prioritize sexuality-related needs (U) |
| Lack of organisational support | Lack of time and resources (U) |
|  | Heavy workload prevents staff from addressing intimacy (U) |
|  | Need for support from the agency (U) |
|  | Lack of policy (U) |
|  | Lack of support (U) |
|  | The need for support from both direct peers and colleagues (U) |
| The need for improved guidelines around intimacy needs | Introduction of guidelines will be welcomed by patients (U) |
|  | Acknowledging service users' need for privacy and respect (U) |
|  | Rigid guidelines are unfavoured by patients (U) |
|  | Desire for guidance and training (U) |
|  | The need for peer-developed guidelines (U) |
|  | Conversations about sexuality and sexual health need to be included in a structured way in forensic psychiatric care (U) |
|  | Making sexual health a part of routine enquiry would support conversations around the topic (E) |
|  | There were no clear guidelines regarding how to systematically address sexuality in everyday hospital life (U) |
|  | Staff felt more comfortable with having a standardised way of asking questions regarding sex and sexuality (U) |
